# Integrating genetic data in target trial emulations improves their design and informs the value of polygenic scores for prognostic and predictive enrichment

**DOI:** 10.1101/2024.11.05.24316763

**Authors:** Jakob German, Zhiyu Yang, Sarah Urbut, Pekka Vartiainen, FinnGen, Pradeep Natarajan, Elisabetta Pattorno, Zoltan Kutalik, Anthony Philippakis, Andrea Ganna

## Abstract

Randomized controlled trials (RCTs) are the gold standard for evaluating the efficacy and safety of medical interventions but ethical, practical, and financial limitations often necessitate decisions based on observational data. The increasing volume of such data has prompted regulatory bodies to rely more on real-world evidence, primarily obtained through trial emulations. This study explores how genetic data can improve the design of both emulated and traditional trials. We successfully emulated four major cardiometabolic RCTs within FinnGen (N=425 483) and showed how reduced differences in polygenic scores (PGS) between trial arms track improved study design and consequently reduced residual confounding. Complementing these results with simulations, we show that PGS cannot be directly used to adjust for residual or unmeasured confounding. Instead, we propose an approach that uses genetic instruments for confounding detection and apply this approach to identify likely confounders in Empareg trial emulation. Finally, our results suggest that trial emulations can inform the practical application of PGS in RCTs, potentially improving statistical power. Such prognostic enrichment strategies need to be assessed in a trial-relevant population, and we show that, for 2 out of 4 emulated trials, the association between PGS and trial outcomes in the general population was different from what observed in the population included in the trial.

In conclusion, our work shows that genetic information can improve the design of emulated trials. These results contribute to the establishment of a promising new era of genetically-informed clinical trials.

## Introduction

When they are available, randomized controlled trials (RCT) are the gold standard to evaluate the comparative efficacy and safety of medical interventions.^1^ Randomization ensures that the interventional and non-interventional groups are closely comparable in their characteristics, thus allowing any observed effects to be causally linked to the treatment under investigation. In many real-world scenarios, however, RCT data are not available, and decisions need to be made based on the data at hand.

As the volume of observational data continues to grow exponentially, regulatory bodies such as the U.S. Food and Drug Administration (FDA) or the European Medicines Agency (EMA) are increasingly inclined to utilize real-world evidence to gain insights into the effectiveness of medical interventions in clinical practice.^2,3^ Trial emulations based on real-world datasets are being increasingly leveraged to this purpose, with ongoing attempts to compare their results with findings from RCTs.^4–6^ However, trial emulations can be biased, and traditional epidemiological limitations of observational analyses, including the exchangeability assumption (“no unmeasured confounding”) remain.^7–9^ Residual and unmeasured confounding pose potential threats to the validity of epidemiological studies.^10^

Trial emulations are typically based on claims or registry data that have detailed information on drug prescription and, importantly, purchases, ensuring accurate tracking of patient medication use. These datasets are large but not deep. They do not capture comprehensive biological information such as genomics and proteomics. Biobank studies, on the contrary, are rich of -omics information, but so far, there have been limited efforts to emulate trials within biobanks.^11,12^ The main reasons are the small sample size and the difficulty to link them with claims data, especially in the US.

Yet, integrating genetic data, alongside comprehensive registry information and expert knowledge, offers a distinctive opportunity to improve trial emulation. For example, genetics offers the opportunity to augment clinical trial design by identifying individuals based on higher risk of disease (‘prognostic enrichment’), or increased probability of benefit (‘predictive enrichment’).^13^ Further exploration of this concept within a trial emulation setting could pave the way for its implementation in subsequent RCTs. For example, trial emulations can be used to understand if polygenic scores can be used for prognostic enrichment within a study population selected with similar inclusion and exclusion criteria as for the RCT, rather than in the general population, as routinely done.^14^

Genetic information is also unique when compared to data available in claims datasets. Genetic information is stable across life, it is not impacted by reverse causation and has low measurement errors. Thousands of genetic variants have been associated with almost every possible measurable human trait creating a unique catalog of genotype-phenotype relationships. Analogously to the common use of e.g. socioeconomic or behavioral indicators as proxy variables for unmeasured confounders, using polygenic scores (PGS) as proxy measures for unobserved variables might represents an opportunity to overcome the challenge of accounting for confounding variables that are absent from the dataset.^15–17^

Moreover, genetic differences among treatment groups in an emulated trial could potentially offer insights into residual confounding effects. Utilizing genetic variants as instrumental variables in a Mendelian Randomization (MR) analysis^18,19^ can help to understand the effect of a potential confounder on the treatment, as well as on the trial outcome at different stages of the emulation process. Genetic information is thus an attractive tool for causal inference and can be used, similar to what has been suggested for other causal inference approaches ^17,20^, to identify unmeasured confounding risks.

In this study, we emulate four cardiometabolic RCTs within FinnGen^21^, a Finnish biobank-based study including 425 483 individuals with extensive linkage to drug purchases and other health records data. Leveraging both real data and simulations, we propose new applications of genetics to detect and mitigate confounding risks in trial emulations. Finally, we show how trial emulations within biobanks can inform on the value of PGS for prognostic and predictive enrichment in RCT.

## Results

### Successful emulation of four major cardiometabolic RCTs in FinnGen

We consider four large cardiometabolic RCTs, two (EMPA-REG OUTCOME^22^ [Empareg], and TECOS^23^ [Tecos]) focused on type 2 diabetes (T2D) patients and two (ARISTOTLE^24^ [Aristotle] and ROCKET-AF^25^ [Rocket]) on patients with atrial fibrillation (AF). Briefly, Empareg established that empagliflozin, a SGLT-2 inhibitor, was associated with a significantly lower risk of cardiovascular events, represented by the composite endpoint 3-point Major Adverse Cardiovascular Events (3P-MACE). Tecos demonstrated that sitagliptin, a DPP4-inhibitor, was non-inferior to usual care for T2D without sitagliptin, with no significant difference in cardiovascular outcomes, as measured by 3P-MACE, thereby confirming the null hypothesis. Aristotle showed that AF patients at increased risk for stroke using apixaban had lower risk of stroke or systemic embolism compared to warfarin users. Among a similar patient population, Rocket showed lower risk of stroke or systemic embolism among rivaroxaban versus warfarin use.

We closely replicated these four RCTs in FinnGen, a Finnish biobank study, using the trial emulation framework (**Figure 1**) used by the RCT-DUPLICATE initiative^26^, a major trial replication initiative that systematically evaluates the feasibility of using real-world evidence to emulate RCTs and assess the concordance of their findings. Patient characteristics for each trial can be found in the **Supplementary Table 1**. **Supplementary Figure 1** and **Supplementary Tables 2-3** contain study design and event rate comparisons between the original RCTs and our emulations. On average, the number of individuals included in the emulated RCTs was smaller than the original trials, with reductions ranging from 36% in the EMPA-REG trial to lower percentages in others, reflecting the large sample sizes typically required for such studies. Despite a considerable number of individuals meeting the inclusion and exclusion criteria, a substantial drop in sample size occurred during 1:1 propensity score nearest-neighbor matching (e.g., N=13,677 eligible individuals in EMPA-REG, reduced to N=4,522 after matching).

**Figure 1.**
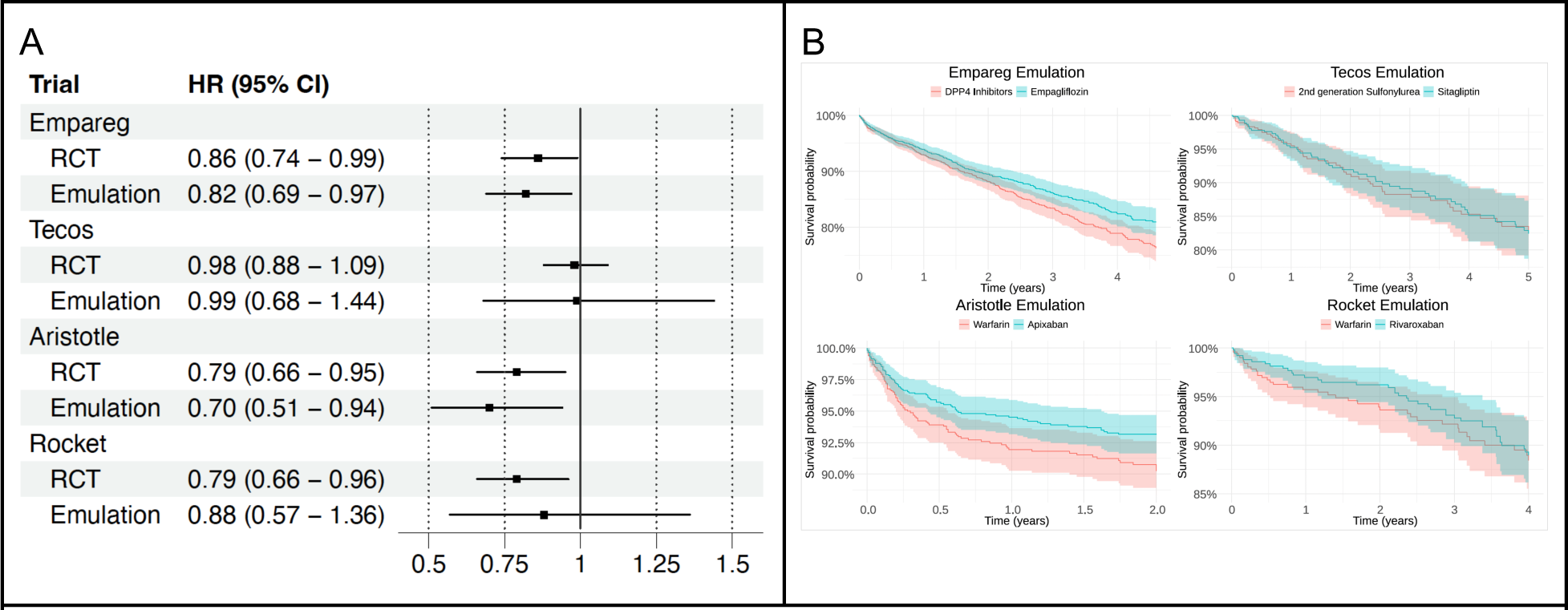
Agreement between randomized controlled trials and their real-world data emulations in FinnGen. A. Comparison of the estimates (HR and 95% CI) of the RCT and their emulation in FinnGen. Empareg (EMPA-REG OUTCOME), BI 10773 (Empagliflozin) Cardiovascular Outcome Event Trial in Type 2 Diabetes Mellitus Patients; Tecos (TECOS), Sitagliptin Cardiovascular Outcomes Study (MK-0431-082); Aristotle (ARISTOTLE), Apixaban for the Prevention of Stroke in Subjects With Atrial Fibrillation; Rocket (ROCKET-AF), An Efficacy and Safety Study of Rivaroxaban With Warfarin for the Prevention of Stroke and Non-Central Nervous System Systemic Embolism in Patients With Non-Valvular Atrial Fibrillation B. Cumulative event Kaplan-Meier plots for primary endpoints in FinnGen trial emulations. HR, hazard ratio; CI, confidence interval; RCT, randomized controlled trial.

For all four emulated RCTs the hazard ratio estimates were within the 95% CI of the original RCT’s estimate and aligned with the same direction of the effect. Thus, according to this definition, and similar to what was done by the RCT-DUPLICATE initiative, all four trials were “successfully” emulated. However, in Rocket, Rivaroxaban was not significantly associated with a lower risk of the composite endpoint stroke/systemic embolism compared to Warfarin (HR = 0.88; 95% CI= 0.57-1.36) whereas the original trial observed a significant risk reduction (HR = 0.79; 95% CI= 0.66-0.96).

### Differences in polygenic scores between trial arms capture emulated trials reduction in residual confounding compared to naïve approaches

Having emulated 4 RCTs in FinnGen, we assess whether genetic information could be used to evaluate the robustness of the emulation approach with regards to confounding

As observational data is not randomized, confounding by indication is a major challenge in observational studies of medications. It occurs when the condition that prompts the prescription of a drug is the true cause of the outcome being studied. For instance, doctors may choose a specific drug based on patient characteristics (such as the severity of the disease or potential for adverse reactions), which are not always fully captured in the data. These characteristics can influence the outcome independently of the medication itself. As a result, differences in outcomes between patients on different drugs may be due to underlying differences in patient characteristics, rather than the effects of the drugs.

To alleviate this bias, emulated RCTs employ a series of precautions, from choosing a sensible comparator group, closely mimicking the trial outcome definition to matching individuals for potential confounders.^27^ However, not all factors considered when prescribing a drug over a comparator are captured in the data. For example, claims data are often poor in capturing laboratory markers. However, genetic information can be used to proxy, albeit imprecisely, many of these biological traits that are not available in observational data.

With this goal in mind, we computed PGSs for 20 traits relevant to cardiometabolic diseases that might capture potential confounders. Some of these traits (e.g coronary heart disease) are directly available in the observational data, and thus matched upon in the emulated trial, others (e.g. C-reactive protein) are not available, as FinnGen currently does not contain information on lab measurements. We examined the genetic differences between the trial arms across different stages of the emulation process with the expectation that, by implementing increasing precautions against bias, the differences in genetically-inferred factors between the trial arms would reduce. Overall, we observed a decreasing trend in genetic differences the higher the level of confounder adjustment (**Figure 2** for Empareg and **Supplementary Figures 2-4** for the other RCTs). In Empareg, we saw a higher imbalance across all PGS in the plain observational setting comparing empagliflozin with non-initiators, which reflects the original RCT design (Empareg vs placebo). We see a particularly high imbalance in the genetically-predicted T2D (standardized mean differences (SMD) = 0.56; 95% CI = 0.54 - 0.57), glycated hemoglobin (HbA1c) (SMD = 0.31; 95% CI = 0.30 - 0.33) and BMI (SMD = 0.21; 95% CI = 0.19 - 0.22) reflecting characteristics of the patient population using empagliflozin. After applying eligibility criteria and considering a sensible comparator group (DPP4 inhibitors users) instead of non-initiators, the PGS differences were overall reduced, but for 7 out of 20 PGS remained statistically significant different between the two arms at a P-value < 2.5 x 10^-3^, including for coronary heart disease (SMD = 0.12; 95% CI= 0.08 - 0.15) and T2D (SMD = 0.08; 95% CI= 0.04 - 0.12). Of note, only T2D patients were included in the RCT emulation stage. Thus, the remaining difference in genetically-predicted T2D likely reflects the difference in liability or risk for T2D between the two arms, which can simply be captured by T2D diagnostic codes.

**Figure 2.**
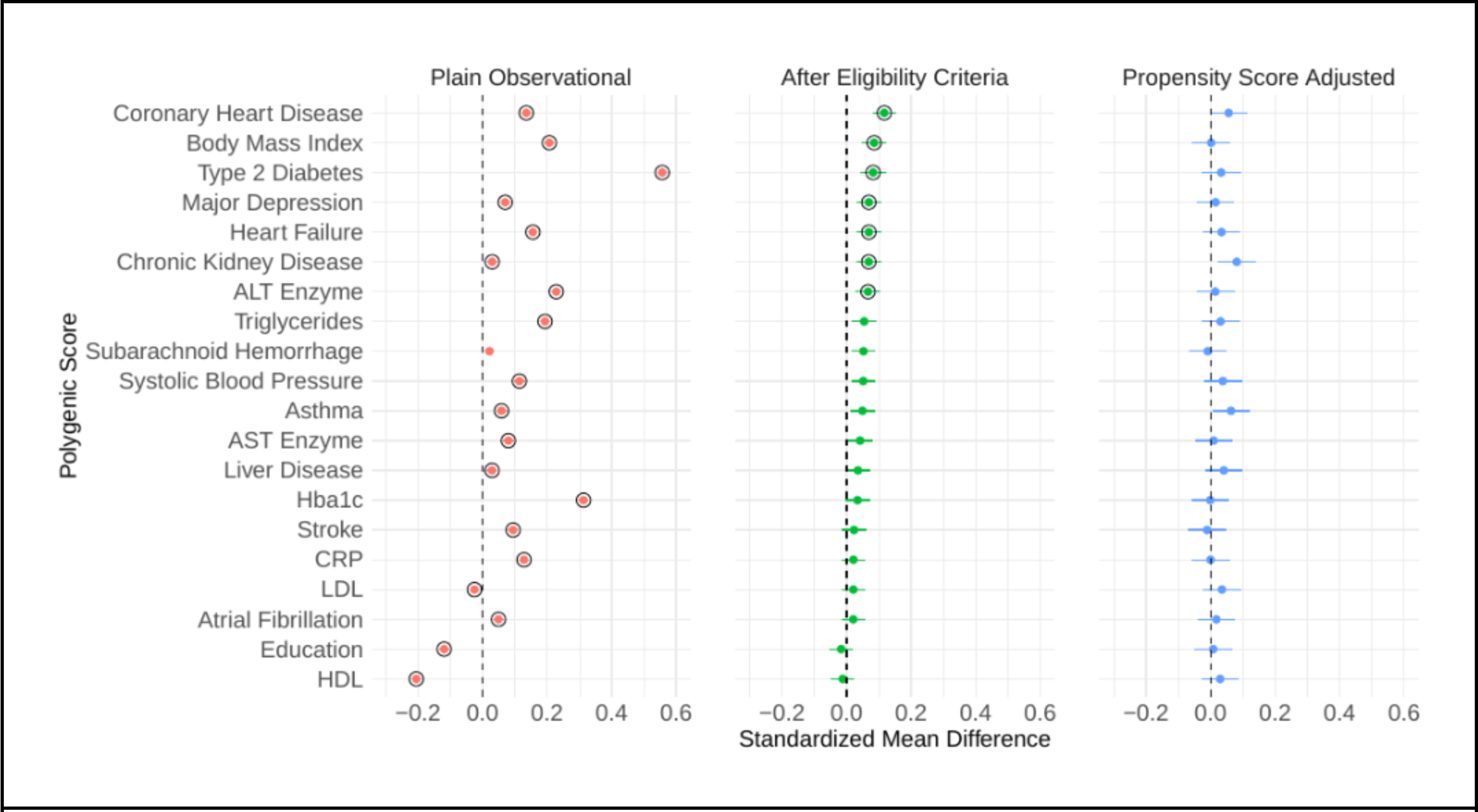
Standardized mean difference of 20 polygenic scores across different stages of the Empareg trial emulation. Plain Observational: empagliflozin initiators vs non-initiators; After Eligibility Criteria: Empareg trial emulation cohort after applying inclusion/exclusion criteria and including an active comparator group (DPP4 inhibitors user). The comparison is between empagliflozin initiators vs DDP4 initiators; Propensity Score Adjusted: Empareg trial emulation cohorts after with inclusion/exclusion criteria and a 1:1 propensity score nearest-neighbor matching for 28 covariates. The comparison is between empagliflozin initiators vs DDP4 initiators. The standardized differences in means of the two trial arms are plotted as point estimates and lines representing their 95% CI. A circle around the point estimates represents statistical significance after a Bonferroni-corrected P value threshold (2.5 x 10^-3^). Analogous plots for the other trial emulations can be found in the **Supplementary Figures 2-4** ALT, alanine transaminase; AST, aspartate transaminase; Hba1c, glycated hemoglobin A1c; CRP, C-reactive protein; LDL, low-density lipoprotein; HDL, high-density lipoprotein; CI, confidence interval.

After 1:1 propensity score nearest-neighbor matching for 26 to 30 covariates, differences were further reduced, and none was significantly different at a P-value < 2.5 x 10^-3^.

For the other three emulated RCTs, we observed similar trends (**Supplementary Figures 2-4)**. Larger PGS differences in the plain observational analysis were observed for non-active comparator RCT (Tecos) vs active-comparator RCTs (Aristotle and Rocket).

### Polygenic scores are unlikely to help controlling for confounding in emulated trials

Having established that PGS differences between trial arms track the level of confounder adjustment, one might speculate that directly controlling for PGS in an emulated trial, for example via propensity score-matching, can help reduce confounders for traits that have not been directly measured.

To better understand this scenario, we constructed directed acyclic graphs^28^ (DAG) and performed simulation studies. The DAG in **Figure 3A** lays out the graphical relationship between treatment, outcome, confounder and PGS assuming PGS is directly causal only to the confounder. Similar to other approaches that use proxy measures for unobserved confounding adjustment^17^, if the PGS was a strongly-predictive causal instrument for the confounder, one might consider adjusting for PGS when the confounder is not available.

**Figure 3.**
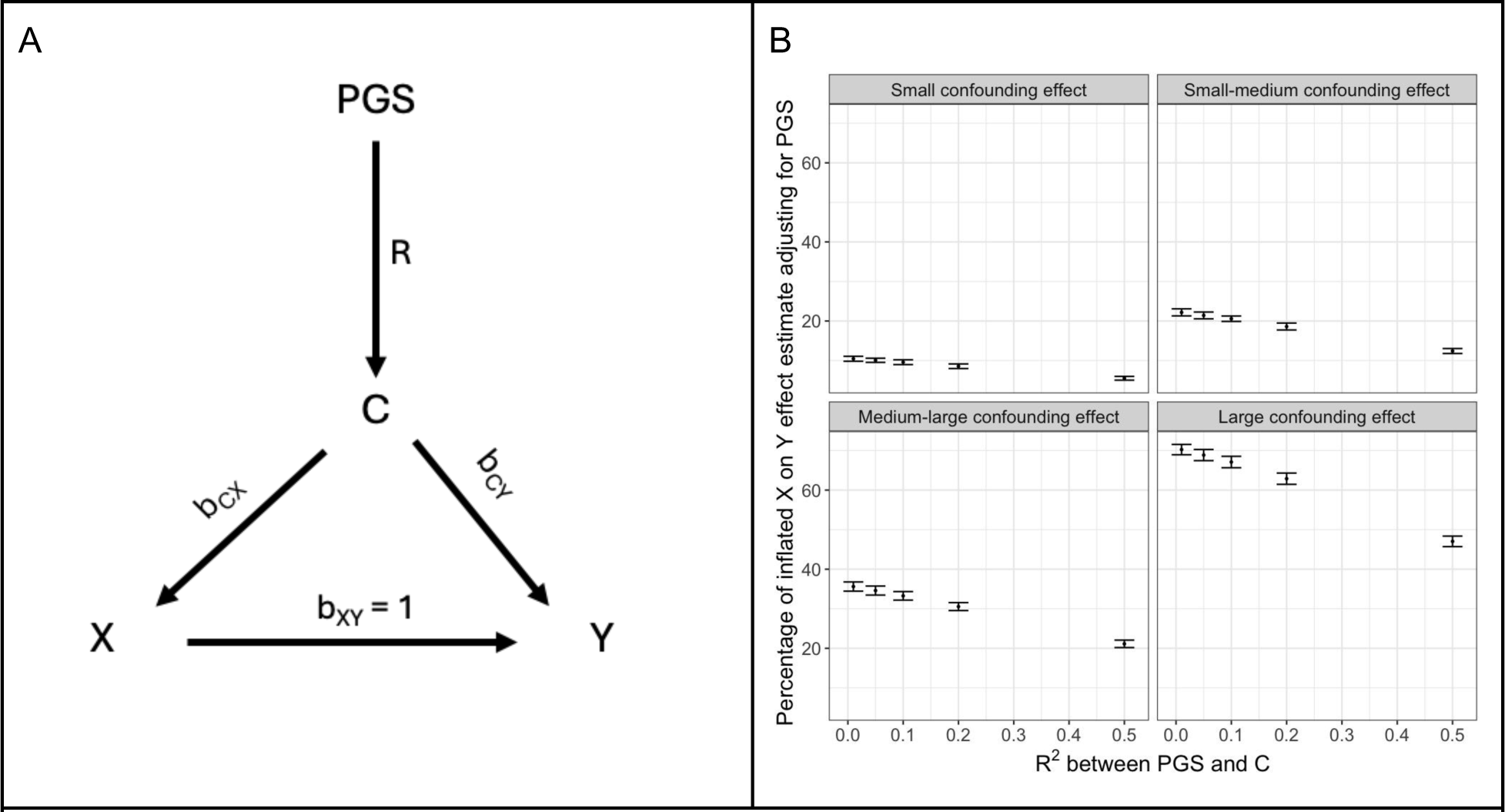
Evaluating the utility of polygenic scores for confounder adjustment. A. A directed acyclic graph (DAG) illustrates the causal structure between polygenic score (PGS), confounder (C), treatment (X) and outcome (Y). The PGS serves as an imperfect proxy variable for the confounder. The effect of the C on the exposure (X) and outcome (Y) are denoted as b_cx_ and b_cy_, respectively. The true unconfounded effect of X on Y is b_xy_ = 1. B. Simulation study: Under this model (see **Methods**), we changed the correlation between C and PGS simply by varying *r,* and the effect of confounding factor C on X and Y by varying b_cx_ and b_cy_. Under each condition, we measured the observed effect of X on Y, conditioned on PGS and calculated the bias as a percentage of the inflated effect of X on Y. 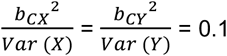 for small confounding effect, 0.2 for small-medium confounding effect, 0.3 for medium-large confounding effect and 0.5 for large confounding effect. These simulations show that even if PGS is strongly correlated with the confounder (i.e. r^2^ = 0.5) -an unlikely scenario given the correlation between PGS and traits are generally lower – correcting for PGS does not completely account for the bias introduced by the confounder.

However, several aspects do not support this claim. The first observation is that while PGS is constructed to predict the confounder it can still be associated with both treatment and/or outcome independent from the confounder. This is because the PGS is a weighted sum of the effects of multiple genetic variants, some of which can be associated with treatment and/or outcome independently of their effect on the confounder (horizontal pleiotropy). We illustrate this possibility with the DAG and simulations in **Supplementary Figure 5**. Thus, controlling for PGS might induce bias by controlling for other non-confounding factors, including mediators.

The second observation is that PGS are generally weak predictors of traits and diseases.^29,30^ Thus, adjusting for PGS would only adjust for part of the variability in the confounders. Under realistic correlation between PGS and the confounder (r^2^ between 0.01 and 0.5) and different magnitude of confounding effect, PGS alone is unlikely to be able to adjust for residual confounding (**Figure 3B** and **Supplementary Figure 6**).

### Mendelian Randomization can help identify residual confounders in emulated trials

Mendelian randomization (MR) is a powerful method to investigate causal relationships between exposure and outcome variables. By leveraging genetic variants as instrumental variables, MR can help infer causality in observational studies.^19,31^

While MR is typically used to assess the causal relationships between an exposure and an outcome, it can be more generally used as a confounder detector.^32^ In this case, genetic variants are used as instruments to test the causal relationships between the potential confounder and both the exposure and the outcome. Unlike PGS, MR selects for variants that are directly associated with the confounder and use different techniques to limit horizontal pleiotropy (i.e. to limit the impact of variants that are associated with the outcome not via the exposure)

We use an MR framework to better understand whether 19 traits can be considered as confounders in the Empareg emulated RCT. Following the DAG in Figure 4A we tested whether the genetic instruments for the potential confounders were associated with both empagliflozin treatment (G → X) and coronary heart disease, a proxy for 3P-MACE (G → Y).

**Figure 4.**
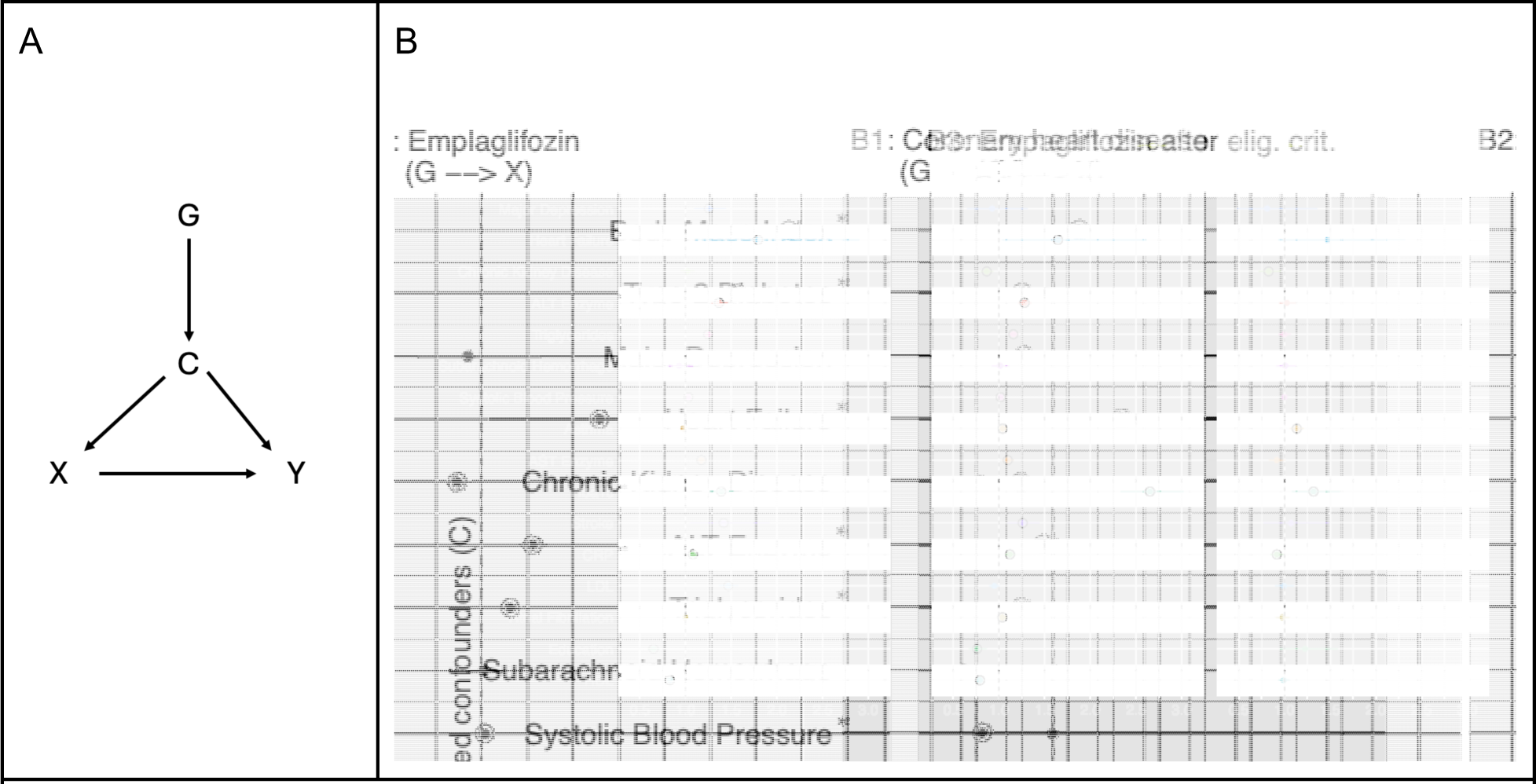
Using mendelian randomization within Empareg trial emulation to identify confounders. A. A direct acyclic graph (DAG) illustrating the relationship between treatment initiation (X) and trial outcome (Y), as well as the effect of a genetic instrument (G) of a confounding variable (C) on both the treatment initiation and trial outcome only through the confounding variable. B. Results of a mendelian randomization (MR) analysis using inverse variance-weighted to study the causal effects of 18 traits on coronary heart disease, representing the trial outcome, and empagliflozin, representing the treatment initiation. B1. MR for association between 18 traits on coronary artery disease using two-sample MR. B2. MR for association between 18 traits on empagliflozin initiation in the full study population. B3 MR for association between 18 traits on empagliflozin initiation after applying the randomized controlled trial’s eligibility criteria. The point estimates represent the odds ratios with lines representing their 95% confidence interval. For continuous confounders, the odds ratio reflects the change in the outcome variable associated with a 1 SD increase in the exposure variable; for binary confounders, the odds ratio represents the change in the outcome variable when comparing the presence versus the absence of the binary exposure. A circle around the point estimates represents statistical significance with a P value threshold of 5 x 10^-2^. * putative confounder, due to significance in B1 and B2; ** putative confounder, due to significance in B1 and B3; ALT, alanine transaminase; AST, aspartate transaminase; Hba1c, glycated hemoglobin A1c; CRP, C-reactive protein; LDL, low-density lipoprotein; HDL, high-density lipoprotein; elig. crit., eligibility criteria; X, treatment; Y, outcome; C, confounder; G, genetic instrument.

Two-sample MR studies revealed putative causal effects of 14 out of 19 potential confounders on coronary heart disease (**Figure 4B1**). There is extensive orthogonal evidence supporting the causal nature of these relationships.^33–38^ When performing MR of the confounder on empagliflozin treatment, we observed 15 out of 19 traits to have a statistically significant effect (**Figure 4B2**). Since confounders are defined as variables with an effect on both, the exposure and outcome, we were specifically interested in traits where we observed an effect on both coronary heart disease and an empagliflozin treatment. This was the case for 12 traits when emulating Empareg with a plain observational approach. For example, BMI was a likely confounder being putatively causally associated, according to MR, with both empagliflozin treatment (OR= 2.68 [2.51 - 2.87], P < 2 x 10^-16^) and coronary heart disease (OR = 1.55 [1.48 - 1.64], P < 2 x 10^-16^). The putative causal effect on empagliflozin treatment highlights doctors’ tendency to prescribe this medication to patients with higher BMI, a significant risk factor for T2D, which is the primary reason for the drug’s prescription.

After including eligibility criteria and a comparator group (**Figure 4B3**), only 2 traits, HbA1c and CRP remain significantly associated, according to MR, to both empagliflozin treatment and coronary heart disease.

We further examined whether the causal effects of potential confounders on empagliflozin treatment was mediated by their effect on coronary heart disease. In other words, if the doctor’s choice to prescribe empagliflozin was informed by the potential confounder effect on the cardiovascular risk of the patient. If that would be the case, the confounder cannot be defined as such as it is associated with exposure via the outcome (C → Y → X). We show that these effects are small across all the putative confounders (**Supplementary Figure 7**) and hence the observed C → Y causal effect is direct.

### Emulated trials can be used to better evaluate the prognostic and predictive enrichment of polygenic scores

PGS can be used to enrich RCTs by identifying individuals based on higher risk of disease (‘prognostic enrichment’), or increased probability of benefit (‘predictive enrichment’).^13^ However, to evaluate these potential benefits, it is necessary to test both prognostic and predictive enrichment hypotheses between a study population that is as close as possible to that of the prospective RCT. In fact, PGS have shown different prediction performances across ages, sex, socio-economic group and co-morbidities.^14,39^ Moreover, eligibility criteria can restrict the study population to high-risk individuals where PGS might have limited effects.^40^

We first test prognostic enrichment by evaluating whether the PGS for the outcomes of the 4 emulated trials (i.e. coronary heart disease and stroke) were associated with the trial outcome within the emulated RCT population (**Figure 5A**). We also compare these effects with those observed in the general population, to see if the performances of PGSs were different when restricting to eligible individuals in the RCTs.

**Figure 5.**
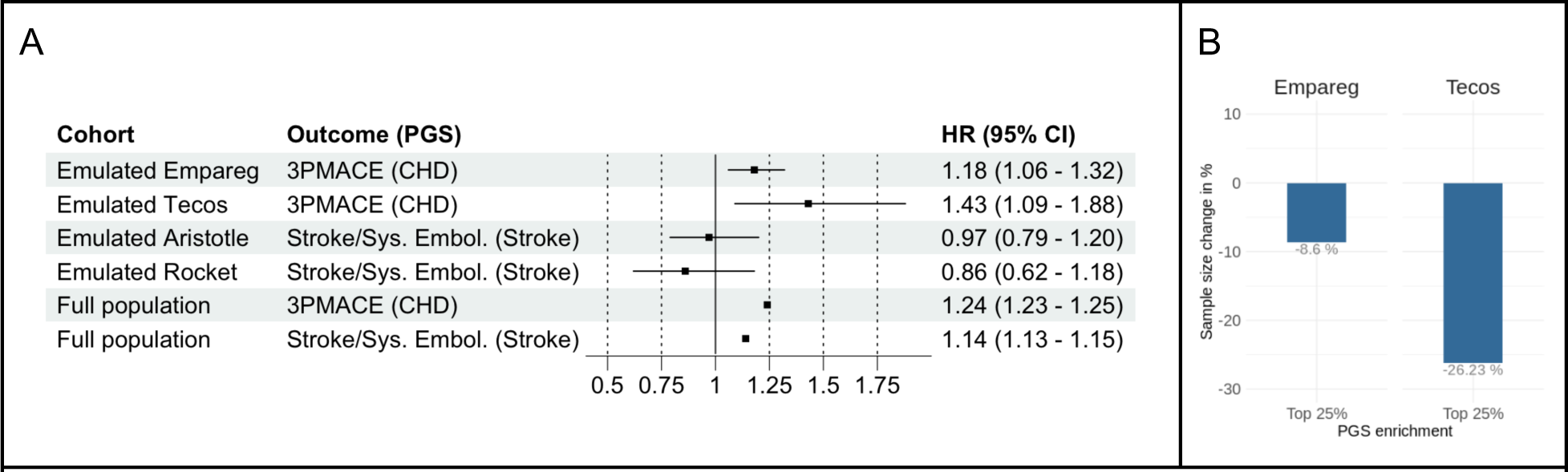
Effect of the polygenic scores on the primary trial outcomes among individuals included in trial emulations and in the full study population. A. Effect of the outcome PGS on the primary outcome within each 1:1 nearest neighbor propensity score-matched trial cohort using Cox regression and adjusting for the treatment, as well as within the full FinnGen population. Hazard ratios per one standard deviation increase in genetic liability and their 95% confidence intervals are illustrated in the central forestplot.B. Sample size reduction of the emulated Empareg and Tecos trials after enriching the trial cohorts with individuals at top 25% genetic risk for CHD (top 25% CHD PGS) Empareg (EMPA-REG OUTCOME), BI 10773 (Empagliflozin) Cardiovascular Outcome Event Trial in Type 2 Diabetes Mellitus Patients; Tecos (TECOS), Sitagliptin Cardiovascular Outcomes Study (MK-0431-082); Aristotle (ARISTOTLE), Apixaban for the Prevention of Stroke in Subjects With Atrial Fibrillation; Rocket (ROCKET-AF), An Efficacy and Safety Study of Rivaroxaban With Warfarin for the Prevention of Stroke and Non-Central Nervous System Systemic Embolism in Patients With Non-Valvular Atrial Fibrillation HR, hazard ratio; CI, confidence interval; PGS, polygenic score, 3PMACE, 3-point major adverse cardiovascular events; CHD, coronary heart disease; Sys. Embol., systemic embolism.

For both Empareg and Tecos emulation, we found the PGS for coronary heart disease to be associated with 3P-MACE (HR = 1.18; 95% CI= 1.06-1.32 in Empareg and HR = 1.43; 95% CI= 1.09-1.88 in Tecos). These effects were consistent with what was observed in the full FinnGen (HR = 1.24; 95% CI= 1.23-1.25).

However, for Aristotle and Rocket emulation the PGS for stroke was not associated with the composite endpoint stroke/systemic embolism (HR = 0.97; 95% CI= 0.79-1.20 for Aristotle and HR = 0.86; 95% CI= 0.62-1.18 for Rocket) despite the significant PGS association in the full population of FinnGen (HR = 1.14; 95% CI= 1.13-1.15).

These results suggest that care should be taken when generalizing the PGS association from the general population to RCT participants.

Given the significant prognostic enrichment for Empareg and Tecos, we calculated the reduction in sample size required to achieve a similar number of events, and consequently similar statistical power, if we had included individuals in the top 25% of the PGS for coronary heart disease. This prognostic enrichment approach strategy would have resulted in −8.6% and −26% reduction in sample size, given all the other inclusion and exclusion criteria being the same (**Figure 5B**).

Finally, we tested predictive enrichment in Empareg and Tecos by evaluating the interaction between the PGS for coronary heart disease and the treatment arm indicator. A significant interaction would indicate the treatment being more effective in individuals with higher or lower PGS. There was no significant interaction either in the Empareg emulation (P = 0.99) or in the Tecos emulation (P = 0.24).

## Discussion

In this study we show how genetic data can benefit target trial emulation design and analysis and how a trial emulation framework can be used to better understand the value of polygenic scores for RCT design.

To answer these questions, we first emulated 4 transformative cardiometabolic RCTs in FinnGen using the framework used by the RCT-DUPLICATE initiative.^26^ We learned that despite the large sample size and complete national coverage of drug purchases and health outcomes, RCT emulation requires a very large number of individuals. During the propensity score matching step, on average, 78% of individuals get discarded, as we aimed to match patients as closely as possible to ensure comparability between treatment groups, thus reducing the final sample size. Nonetheless, we were able to successfully emulate all 4 trials and generate real-world evidence that is concordant with the RCTs results. Emulation of smaller RCTs, as e.g. trials for rare diseases, is probably not possible at the current sample size of 500,000 genotyped individuals, highlighting how generation of ever larger genetic datasets is required if one aims to assess the role of genetics in trial-like populations.

Confounding by indication is particularly severe in observational studies of medications. Our approach leverages the enormous catalog of genotype-phenotype relationships generated by genome-wide association studies to “impute” biological risk factors that might act as confounders. We show that polygenic scores can be used to identify both measured and unmeasured factors that are unbalanced between the two arms of the trials. Some of these factors are likely confounders, others are not; polygenic scores cannot distinguish between the two. However, polygenic scores can provide a more refined measure of the disease risk than simple disease diagnoses. For example, we show that in the emulated EMPAREG trial, which includes only T2D patients, a polygenic score with T2D was still unbalanced between the two arms of the trial. This might reflect unaccounted confounding by indication based on patients’ T2D risk of T2D-related factors. Reassuringly, we saw that in all emulated trials, the polygenic scores imbalance greatly reduced after propensity score matching. Our work highlights the importance of matching as a technique for confounding adjustment in observational data and suggest that polygenic scores can be used as an orthogonal assessment of the quality of matching, especially for biologically risk factors with genetic bases that are not comprehensively captured by claim or registry data (e.g. disease-specific biomarkers). It is also worth highlighting that if a polygenic score is balanced between trial arms, this does not imply the predicted trait is also balanced. Polygenic scores are generally poor predictors of traits, even those with strong genetic bases, and adjusting or matching for polygenic scores, as shown by our simulations, is unlikely to control for the trait they are predicting. One should also consider that if the polygenic score for a potential confounder correlates with the genetics of the drug response, PGS differences between trial arms should be expected.

Genetics-based instrumental variable approaches can however be used, together with specialist knowledge and other orthogonal evidence, to identify confounders. While others suggested that Mendelian Randomization can be used for confounder detection ^41–43^, we applied and extended this framework to emulated RCTs. While MR has numerous limitations that have been extensively described with regard to its most common use to assess the causal relationships between exposure and outcome^19^, here we mention a few limitations that are unique to its use in confounder detection. First, the causal relationship between an exposure (or potential confounder) and a treatment should be interpreted with caution. A putative causal effect is likely to indicate that the exposure is influencing the doctor’s decision to prescribe the treatment and not the treatment effectiveness itself. For example, we identify a negative putative causal effect of chronic kidney disease (CKD) on empagliflozin treatment, reflecting the former doctor’s decision to avoid prescribing empagliflozin to patients with severely impaired renal function before more recent evidence emerged showing its benefits for these patients.^44–47^ Second, the effect of the confounder on the treatment can be mediated by the outcome or the effect on the outcome can be mediated by the treatment. These effects can be addressed by closer examining the true causal structure and adjusting the confounder effects by the effects of all other pathways, excluding the direct effect of confounder on treatment or outcome, respectively. Despite these interpretational challenges, MR can be a powerful tool for confounder detection during RCT emulation. At the “eligibility criteria” stage MR could inform about residual confounding and suggest which factors, if measured, to include in propensity score-matching. After matching, MR can still be used to investigate non-adjusted residual confounding and, together with expert knowledge, better interpret the results of the emulated RCTs. While we only discussed MR for confounding detection, it would be theoretically possible to use MR for confounding adjustment.

The trial emulation framework is useful to better understand the value of genetics in trial design. The most promising use of trial emulation is to assess the prognostic enrichment for polygenic scores. Individuals enrolled in RCTs are highly selected and do not represent the general population. It would be naïve to assume the magnitude of association of a polygenic score with a certain outcome in the general population would be the same among RCT trial participants. A trial emulation framework can be used to draw a boundary on the expected association between the polygenic score and the trial outcome, a key piece of information when designing a RCT that uses genetics for either patient selection or as stratification criteria.

A trial emulation is less valuable to understand predictive enrichment because, at current sample size, biobank-based emulated RCTs still have limited power to test for interaction between polygenic scores and treatment or to stratify individuals in different genetic risk bins.

In conclusion, our work shows that genetic information can improve the design of emulated trials, which, in turn, can help inform the use of genetics in designing RCTs. Some of these results can be extended to other -omics that are getting measured in hundreds of thousands of biobanked samples.

## Methods

### Study population

In the current study, we included samples from 425 483 individuals from Finland, sourced from FinnGen Data Freeze 10 (https://www.finngen.fi/en).^21^ This biobank study includes samples from hospital biobanks, alongside prospective epidemiological and disease-based cohorts. Utilizing the unique national personal identification numbers, the data were interconnected with national registries including hospital discharge records (accessible from 1968), death records (from 1969), cancer registries (from 1953), and drug purchase records (from 1995). Registry information was accessible up to December 31, 2021.

### Trial Selection

As the currently largest trial emulation effort, the RCT Duplicate project^6,26^ has been emulating numerous RCTs in US-American insurance claims datasets, the goal of which was to assess the utility of the obtained Real-World Evidence (RWE) for regulatory-decision making.

We sought to identify four RCTs that have been previously replicated by RCT Duplicate and were feasible to be successfully emulated in our Real-World Data (RWD) dataset. By the time of initiation of our project, findings of the first 10 trial emulations were published by the RCT Duplicate project.^26^ The evaluation criteria deciding upon the feasibility of a RCT replication included critical aspects of the trial emulation protocol, such as the primary outcomes, eligibility criteria, treatment strategies, allowing for only minor deviations if features were not available in our data source (**Supplementary Tables 2-3**). The RCT was seen as closely emulated when the emulation of comparator and outcome were at least moderate, and at least one of them was good, as described in the meta-analysis of RCT Duplicate data.^48^

### Trial Emulation Design and Analysis

Based on RCT Duplicate’s trial emulation efforts, we developed the protocols for the emulations of four trials (Empareg, Tecos, Aristotle and Rocket). Closely following the original trial protocols we emulated an observational data protocol for each trial, including the eligibility criteria, treatment strategies, assignment procedures, follow-up periods, primary outcomes, causal contrasts and an analysis plan.^4^

Different sets of eligibility criteria required fulfillment within distinct timeframes prior to therapy initiation. Flowcharts of cohort formations can be found in **Supplementary Tables 4-7** and **Supplementary Figures 8-11**.

The treatment strategies included new users of either the drug of interest or the comparator drug, starting from the date the newer drug received marketing authorization in Finland. For the two placebo-controlled trials, Empareg and Tecos, we selected an active comparator as a proxy for placebo regarding cardiovascular effects, similar to RCT Duplicate. This is due to the fact that confounding bias may become especially serious when active user groups are compared to nonuser groups, as nonuser comparator groups considerably differ from actively treated patients in ways that are poorly captured in observational datasets.^49,50^

As a proxy for placebo DPP4-inhibitors for Empareg and second-generation sulfonylureas for Tecos were chosen, given they likewise antidiabetic treatments, commonly prescribed interchangeably to the treatments of interest and are known not have any causal effect on cardiovascular outcomes based on current evidence.^23,51–53^

As the assignment procedures in observational studies are never at random, an adjustment for confounding variables is required in order to satisfy the exchangeability assumption. We selected sets of >25 confounding variables, measured within 6 months prior to drug initiation, reflecting demographics, comorbidities, comedications and cardiovascular procedures. We adopted 1:1 propensity score (PS) nearest-neighbor matching with a caliper of 0.1 or 0.01 on the PS scale, depending on the initial overlap.^54,55^ PS matching statistics and details on covariate balance for all trial emulations can be found in **Supplementary Tables 8-11**.

Follow-up started at the first purchase of either of the defined therapeutics and ended with the occurrence of a primary outcome event, death, discontinuation or switch to a comparator or end of registry information, whichever occurs first. The time point of a discontinuation of therapy was calculated based on the number of packages purchased by the patient multiplied by the package size.

The primary outcome for Empareg and Tecos was 3P-MACE and for Aristotle and Rocket a composite endpoint of stroke and systemic embolism, adapted from the definition used in the corresponding trials.

In our analysis we employed an “on-treatment” approach attempting to replicate an intention-to-treat estimate derived from the RCT with particularly high treatment compliance. Hazard ratios (HR) and 95% confidence intervals (95% CI) were estimated in PS-matched cohorts using the Cox proportional hazard models. We defined “estimate agreement” as the emulation estimate being within the 95% CI for the RCT estimate.

### PGS Generation

We computed genome-wide polygenic scores (PGS) for 20 traits (**Supplementary Table 12**) using the PGS-continuous shrinkage priors (CS) method.^56^ The input weights were derived from available summary statistics sourced from external GWAS data pertaining to the 20 traits. Variants were restricted to those present in the HapMap 3 reference panel.^57^ To ensure comparability, PGS were standardized (mean = 0; standard deviation = 1) in the whole FinnGen population. Detailed information regarding the summary statistics can be found in the supplementary material.

### PGS Analysis of Cohorts

We investigated genetic differences between the treated and control groups at three different stages of the emulation process and how they change with increased confounder adjustment.

For each PGS we calculated the difference in means (standardized mean difference, SMD) between the treated and control groups using logistic regression and determined its significance on the basis of a Bonferroni-corrected P value threshold (2.5 x 10^-3^).

In the first stage, we looked at a plain observational setting that is best reflecting the original RCT question. Therefore, as Empareg and Tecos are both placebo-controlled trials, we defined the plain observational setting as initiators of the treatment vs non-initiators. Since Aristotle and Rocket are both active-comparator trials, the plain observational setting was defined as initiators of the treatment vs initiators of the active comparator. In the second stage, we looked at the cohorts after applying the eligibility criteria. And in the third, we considered the PS-matched cohorts.

### Simulations

To show that correcting on an imperfect proxy of the confounder can result in bias in effect size estimates, we carried out simulation experiments under the causal model shown in **Figure 3A**. We first generated PGS as a random variable following the standard normal distribution N(0,1), and the rest of the variables were subsequently created as

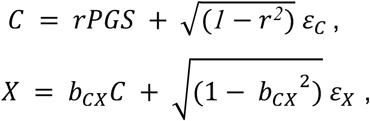

and *Y* = *X* + *b_CY_C*, where *ɛ_C_*, *ɛ_X_ ∼ N*(0, 1).

The variables were simulated as such so that the variance of PGS, C and X were all 1, and the expected effect of X on Y is 1. Under this model, we could change the correlation between C and PGS simply by varying r, and the effect of confounding factor C on X and Y by varying b_CX_ and b_CY_. Under each condition, we measured the observed effect of X on Y, conditioned on PGS, which was an imperfect proxy of C, through linear regression *lm*(*Y* ∼ *X* + *PGS*). We denoted estimate bias as the observed regression coefficient – 1, which is the expected underlying effect of X on Y.

We wanted to also demonstrate that even under a fixed correlation coefficient between C and PGS, extent of bias in observed X on Y effect can still vary due to additional components contributing to only PGS and X, Y but not C, we further carried out simulations under a different causal model showed in **Supplementary Figure 6A**, where PGS and confounder C are correlated due to a common underlying causal factor *G*^∗^. Meanwhile, an extra component G’ contribute only to PGS but not C. Under this model, we first generated shared causal factor *G*^∗^ and PGS unique causal factor G’ independently following the standard normal distribution *N(0,1)*, and other variables as below:

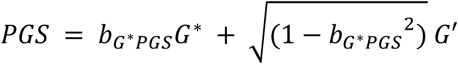

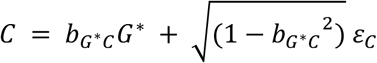 where 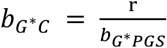 and r is the correlation coefficient between C and PGS. We fixed the contribution of *G*^∗^ on PGS as 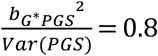 and *r*^2^ *=* 0.3 in this experiment.

Subsequently, we simulated X and Y as

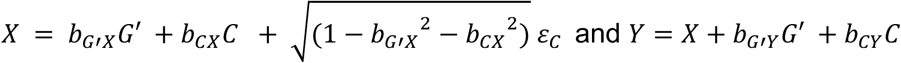

The variables were simulated as such so that variance of G’, G*, PGS, C and X were all 1, and the expected effect of X on Y is 1. In this experiment, for simplicity, we fixed the contribution of C on X and Y so that 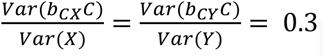, and assumed that G’ has no effect on Y. Furthermore, as a proof of concept, we assumed that G’ has a negative effect on 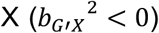 since in this case, we expect to see an increment in estimate bias when G’ contributes more to the variance of X. We looked at estimate bias from a same linear regression *lm*(*Y* ∼ *X* + *PGS*) in respect of changes in 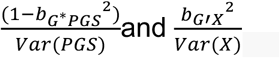.

### Genome-wide association studies

We used REGENIE^58^ to perform a GWAS of empagliflozin initiation in the whole population, including 426,775 samples (cases: 14,996; controls: 411,779) as well as after applying the eligibility criteria of the Empareg emulation, including 11,349 samples (cases: 4,630; controls: 6,719). Details on genotyping and imputation in FinnGen can be found in Kurki et al. 2023.

### Mendelian Randomization Analysis

By utilizing genetic variants as instrumental variables, we employed two-sample Mendelian randomization (MR) to investigate the confounding status of numerous variables in a trial emulation setting.^18^ In our MR analysis we only focused on the Empareg trial emulation. We examined the effect of the 20 traits used in the PGS analysis (sources of external summary statistics can be found in **Supplementary Table 12**) on the trial outcome, represented by summary statistics for CHD, as well as on receiving the empagliflozin treatment in the whole population and after applying the eligibility criteria, both represented by summary statistics from our GWASs. We performed the MR analysis using the inverse variance-weighted method (IVW).

To obtain the independent instrumental variants (IVs) for each trait we filtered for significant exposure-associated SNPs (P Value < 5 × 10^-8^), performed linkage disequilibrium (LD) clumping (r^2^ < 0.001; clumping window = 10,000 kb) and excluded potential outcome-associated SNPs (defined as P Value < 5 × 10^-8^ with the outcome).

We identified three key steps in using MR to explore confounding: (1) MR of potential confounder on treatment. Conducting an MR analysis to assess the causal effect of the proposed confounding trait on the treatment variable. If the MR analysis shows a significant association, it suggests the potential confounder is indeed related to the treatment. (2) MR of potential confounder on outcome. Performing a separate MR analysis to evaluate the causal effect of the proposed confounding trait on the trial outcome variable. If the MR analysis demonstrates a significant association, it indicates the potential confounder is also related to the outcome. (3) Interpretation. If both MR analyses (steps 1 and 2) show significant associations, it implies the proposed trait is very likely to be a true confounder that needs to be accounted for and addressed through statistical adjustment in the trial emulation to obtain widely unbiased average treatment effects. Expert knowledge is still required to assess the plausibility of the MR analyses.

### Statistical Analysis of the Outcome PGS within Trial Emulations

Analogously to the MR analysis, we selected the CHD PGS as outcome PGS for MACE and Stroke PGS for the composite endpoint stroke/systemic embolism. We evaluated the effect of the outcome PGS on the primary outcome within each PS-matched cohort using Cox regression and adjusting for the treatment.

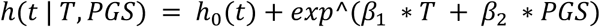

*h*(*t*) : hazard at time t

*h*_0_(*t*) : baseline hazard at time t

*T* : treatment group

*PGS* : outcome PGS

*β*_1_ and *β*_2_ : coefficients associated with the treatment group variable *T* and *PGS* respectively

Additionally, we predicted the outcome PGS effects on the primary outcome in the full population, using Cox regression. Survival times started at birth with follow-up until the occurrence of the primary outcome, death or end of registry information, whichever occurred first.

Furthermore, we determined the event rate of the primary outcome for each trial and investigated the event rates within individuals with top 25% PGS. Based on that we calculated the required sample sizes given the new event rates, to reach the same statistical power. This was in order to assess the effect of PGS enrichment on sample sizes in clinical trials.

## Code Availability

The study utilized previously published analysis tools as described in the **Methods** section. Additional code used for these analyses is available at https://github.com/dsgelab/trial_emulations_genetics

## Data Availability

Access to individual-level sensitive health data, as mandated by National and European regulations (GDPR), requires approval from national authorities for specific research projects and for researchers who are explicitly listed and approved. The health data referenced in this study was generated and provided by the National Health Register Authorities (Finnish Institute of Health and Welfare, Statistics Finland, KELA, Digital and Population Data Services Agency) and approved by either the respective authorities or the Finnish Data Authority, Findata, for use in the FinnGen project. As a result, we, the authors, are unable to grant access to individual-level data to third parties. However, researchers can apply for access to the health register data through the Finnish Data Authority, Findata (https://findata.fi/en/permits/), and for individual-level genotype data from Finnish biobanks through the Fingenious portal (https://site.fingenious.fi/en/), managed by the Finnish Biobank Cooperative FINBB (https://finbb.fi/en/). All Finnish biobanks can provide data for research projects under the scope of the Finnish Biobank Act, which includes research aimed at promoting health, understanding disease mechanisms, or developing health and medical care products and practices. More information on accessing FinnGen data can be found here: https://www.finngen.fi/en/access_results. A comprehensive list of FinnGen endpoints is available at: https://www.finngen.fi/en/researchers/clinical-endpoints.

## Ethics statement and materials & methods

Patients and control subjects in FinnGen provided informed consent for biobank research, based on the Finnish Biobank Act. Alternatively, separate research cohorts, collected prior the Finnish Biobank Act came into effect (in September 2013) and start of FinnGen (August 2017), were collected based on study-specific consents and later transferred to the Finnish biobanks after approval by Fimea (Finnish Medicines Agency), the National Supervisory Authority for Welfare and Health. Recruitment protocols followed the biobank protocols approved by Fimea. The Coordinating Ethics Committee of the Hospital District of Helsinki and Uusimaa (HUS) statement number for the FinnGen study is Nr HUS/990/2017.

The FinnGen study is approved by Finnish Institute for Health and Welfare (permit numbers: THL/2031/6.02.00/2017, THL/1101/5.05.00/2017, THL/341/6.02.00/2018, THL/2222/6.02.00/2018, THL/283/6.02.00/2019, THL/1721/5.05.00/2019 and THL/1524/5.05.00/2020), Digital and population data service agency (permit numbers: VRK43431/2017-3, VRK/6909/2018-3, VRK/4415/2019-3), the Social Insurance Institution (permit numbers: KELA 58/522/2017, KELA 131/522/2018, KELA 70/522/2019, KELA 98/522/2019, KELA 134/522/2019, KELA 138/522/2019, KELA 2/522/2020, KELA 16/522/2020), Findata permit numbers THL/2364/14.02/2020, THL/4055/14.06.00/2020, THL/3433/14.06.00/2020, THL/4432/14.06/2020, THL/5189/14.06/2020, THL/5894/14.06.00/2020, THL/6619/14.06.00/2020, THL/209/14.06.00/2021, THL/688/14.06.00/2021, THL/1284/14.06.00/2021, THL/1965/14.06.00/2021, THL/5546/14.02.00/2020, THL/2658/14.06.00/2021, THL/4235/14.06.00/2021, Statistics Finland (permit numbers: TK-53-1041-17 and TK/143/07.03.00/2020 (earlier TK-53-90-20) TK/1735/07.03.00/2021, TK/3112/07.03.00/2021) and Finnish Registry for Kidney Diseases permission/extract from the meeting minutes on 4th July 2019.

The Biobank Access Decisions for FinnGen samples and data utilized in FinnGen Data Freeze 10 include: THL Biobank BB2017_55, BB2017_111, BB2018_19, BB_2018_34, BB_2018_67, BB2018_71, BB2019_7, BB2019_8, BB2019_26, BB2020_1, BB2021_65, Finnish Red Cross Blood Service Biobank 7.12.2017, Helsinki Biobank HUS/359/2017, HUS/248/2020, HUS/150/2022 § 12, §13, §14, §15, §16, §17, §18, and §23, Auria Biobank AB17-5154 and amendment #1 (August 17 2020) and amendments BB_2021-0140, BB_2021-0156 (August 26 2021, Feb 2 2022), BB_2021-0169, BB_2021-0179, BB_2021-0161, AB20-5926 and amendment #1 (April 23 2020)and it’s modification (Sep 22 2021), Biobank Borealis of Northern Finland_2017_1013, 2021_5010, 2021_5018, 2021_5015, 2021_5023, 2021_5017, 2022_6001, Biobank of Eastern Finland 1186/2018 and amendment 22 § /2020, 53§/2021, 13§/2022, 14§/2022, 15§/2022, Finnish Clinical Biobank Tampere MH0004 and amendments (21.02.2020 & 06.10.2020), §8/2021, §9/2022, §10/2022, §12/2022, §20/2022, §21/2022, §22/2022, §23/2022, Central Finland Biobank 1-2017, and Terveystalo Biobank STB 2018001 and amendment 25th Aug 2020, Finnish Hematological Registry and Clinical Biobank decision 18th June 2021, Arctic biobank P0844: ARC_2021_1001.

## Supporting information

FinnGen authors

## Acknowledgements

This work was supported by funding from the Eric and Wendy Schmidt Center at the Broad Institute of MIT and Harvard.

The FinnGen project receives funding from two Business Finland grants (HUS 4685/31/2016 and UH 4386/31/2016) and the following industry partners: AbbVie Inc., AstraZeneca UK Ltd., Biogen MA Inc., Bristol Myers Squibb (and Celgene Corporation & Celgene International II), Genentech Inc., Merck Sharp & Dohme LLC, Pfizer Inc., GlaxoSmithKline Intellectual Property Development, Sanofi US Services, Maze Therapeutics Inc., Janssen Biotech Inc., Novartis AG, and Boehringer Ingelheim International GmbH. We acknowledge the contributions of the following biobanks for providing samples to FinnGen: Auria Biobank (https://www.auria.fi/biopankki/), THL Biobank (https://www.thl.fi/biobank), Helsinki Biobank (https://www.helsinginbiopankki.fi), Biobank Borealis of Northern Finland (https://www.ppshp.fi/Tutkimus-ja-opetus/Biopankki/Pages/Biobank-Borealis-briefly-in-English.aspx), Finnish Clinical Biobank Tampere (https://www.tays.fi/en-US/Research_and_development/Finnish_Clinical_Biobank_Tampere), Biobank of Eastern Finland (https://www.ita-suomenbiopankki.fi/en), Central Finland Biobank (https://www.ksshp.fi/fi-FI/Potilaalle/Biopankki), Finnish Red Cross Blood Service Biobank (www.veripalvelu.fi/verenluovutus/biopankkitoiminta) and Terveystalo Biobank (https://www.terveystalo.com/fi/Yritystietoa/Terveystalo-Biopankki/Biopankki/). All Finnish biobanks are members of the BBMRI.fi infrastructure (https://www.bbmri.fi). The FINBB (https://finbb.fi/) is the coordinator of BBMRI-ERIC operations in Finland. Access to Finnish biobank data is facilitated through the Fingenious services (https://site.fingenious.fi/en/) operated by FINBB.

## Notes

### Competing Interest Statement

The authors have declared no competing interest.

### Author Declarations

Ethics Committee of the FinnGen study and the participating biobanks gave ethical approval for this work

