## Supplementary Tables for "Integrating genetic data in target trial emulations improves their design and informs the value of polygenic scores for prognostic and predictive enrichment"

**Supplementary Table 1 | Patient characteristics for each emulated trial.**

| | Empareg emulation | | | Tecos emulation | | | Aristotle emulation | | | Rocket emulation | | |
|---|---|---|---|---|---|---|---|---|---|---|---|---|
| | Total | Exposure | Comparator | Total | Exposure | Comparator | Total | Exposure | Comparator | Total | Exposure | Comparator |
| **Age (in years), mean ±SD** | 67.3 ±11.0 | 66.9 ±10.9 | 67.6 ±11.0 | 65.0 ±8.7 | 65.6 ±9.0 | 64.3 ±8.3 | 72.4 ±10.3 | 72.3 ±10.5 | 72.5 ±10.2 | 76.9 ±8.9 | 76.7 ±9.0 | 77.2 ±8.8 |
| **Female, %** | 40.0 | 39.8 | 40.2 | 41.8 | 41.5 | 42.0 | 42.8 | 42.9 | 42.7 | 41.5 | 41.0 | 41.9 |
| **History of MI, %** | 3.3 | 3.6 | 3.0 | 1.5 | 1.1 | 1.8 | 5.8 | 5.8 | 5.7 | 5.2 | 5.6 | 4.7 |
| **Hypertension, %** | 17.8 | 17.2 | 18.4 | 11.2 | 12.2 | 10.1 | 28.4 | 25.5 | 31.2 | 49.1 | 48.5 | 49.7 |
| **Heart Faillure, %** | 12.8 | 12.5 | 13.0 | 8.4 | 6.9 | 9.9 | 24.2 | 24.1 | 24.3 | 37.6 | 35.9 | 39.2 |
| **Diabetes, %** | 100.0 | 100.0 | 100.0 | 100.0 | 100.0 | 100.0 | 22.1 | 21.2 | 22.9 | 33.2 | 33.4 | 33.0 |
| **Previous CVD, %** | 39.5 | 40.0 | 39.0 | 26.3 | 26.7 | 25.8 | 100.0 | 100.0 | 100.0 | 100.0 | 100.0 | 100.0 |
| **Atrial Fibrillation, %** | 10.6 | 10.7 | 10.4 | 3.8 | 4.2 | 3.3 | 100.0 | 100.0 | 100.0 | 100.0 | 100.0 | 100.0 |
| **CKD, %** | 1.1 | 1.0 | 1.2 | 0.6 | 0.7 | 0.5 | 3.1 | 2.9 | 3.2 | 0.5 | 0.6 | 0.4 |
| **Asthma, %** | 6.3 | 6.3 | 6.3 | 3.6 | 3.3 | 3.8 | 7.3 | 7.4 | 7.1 | 9.1 | 9.7 | 8.5 |
| **Use of Statins, %** | 73.5 | 74.0 | 72.9 | 66.5 | 66.7 | 66.2 | 53.7 | 54.2 | 53.1 | 60.4 | 61.0 | 59.8 |

| | |
|---|---|
| SD | Standard deviation |
| MI | Myocardial infarction |
| CVD | Cardiovascular disease |
| CKD | Chronic kidney disease |

**Supplementary Table 2 | Comparison between RCT and FinnGen emulation.**
Comparison of trial design between RCT and FinnGen emulation across all studies with respect to exposure, comparator and outcome.

| | | | | RCT | | | FinnGen Emulation | | |
|---|---|---|---|---|---|---|---|---|---|
| **Trial name** | **NCT** | **Type** | **Clinical Area** | **Exposure** | **Comparator** | **Outcome** | **Exposure** | **Comparator** | **Outcome** |
| EMPA-REG OUTCOME | NCT04215536 | Sup | T2D | Empagliflozin | Placebo | 3P-MACE | Empagliflozin | DPP4 inhibitors | 3P-MACE |
| TECOS | NCT03936062 | NI | T2D | Sitagliptin | Placebo | 3P-MACE | Sitagliptin | 2G Sulfonylureas | 3P-MACE |
| ARISTOTLE | NCT04593030 | NI | DOAC AF | Apixaban | Warfarin | Stroke/Syst. Embol. | Apixaban | Warfarin | Stroke/Syst. Embol. |
| ROCKET-AF | NCT04593056 | NI | DOAC AF | Rivaroxaban | Warfarin | Stroke/Syst. Embol. | Rivaroxaban | Warfarin | Stroke/Syst. Embol. |

| | |
|---|---|
| NCT | National Clinical Trial number |
| RCT | Randomized controlled trial |
| 3P-MACE | Composite of nonfatal stroke, nonfatal myocardial infarction, and cardiovascular death |
| AF | Atrial fibrillation |
| DOAC | Direct oral anticoagulant |
| Sup | Superiority trial |
| NI | Non-inferiority trial |
| T2D | Type 2 diabetes |
| DPP4 | Dipeptidyl peptidase 4 |
| 2G | Second generation |
| Syst. Embol. | Systemic embolism |

**Supplementary Table 3 | Study sizes and events.**
Comparison of study sizes and events between RCT and FinnGen emulation across all studies.

| Study | Outcome | RCT | | | | | | FinnGen Emulation | | | | | |
|---|---|---|---|---|---|---|---|---|---|---|---|---|---|
| | | Exposure | | | Comparator | | | Exposure | | | Comparator | | |
| | | Events | N | Rate* | Events | N | Rate | Events | N | Rate* | Events | N | Rate |
| EMPA-REG OUTCOME | 3P-MACE | 490 | 4687 | 3.7 | 282 | 2333 | 4.4 | 241 | 2261 | 5.18 | 314 | 2261 | 6.26 |
| TECOS | 3P-MACE | 839 | 7257 | 4.1 | 851 | 7266 | 4.2 | 56 | 547 | 3.95 | 53 | 547 | 4.05 |
| ARISTOTLE | Stroke/Syst. Embol. | 212 | 9120 | 1.3 | 265 | 9081 | 1.6 | 73 | 1146 | 3.84 | 94 | 1146 | 6.29 |
| ROCKET-AF | Stroke/Syst. Embol. | 188 | 6958 | 1.7 | 241 | 7004 | 2.2 | 41 | 515 | 2.84 | 39 | 515 | 3.26 |

*Events per 100 person-years

| | |
|---|---|
| RCT | Randomized controlled trial |
| 3P-MACE | Composite of nonfatal stroke, nonfatal myocardial infarction, and cardiovascular death |
| Syst. Embol. | Systemic embolism |

**Supplementary Table 4 | Flowchart of the study cohort assembly for the Empareg trial emulation.**
The table lists the included eligibility criteria and the number of the excluded and remaining patients at each step.

| | **Excluded patients** | **Remaining patients** | **% discarded** |
|---|---|---|---|
| All patients | - | 425 483 | |
| Patients who used exposure or a reference | -392 797 | 32 686 | -92,32 % |
| Excluded due to prior use of referent/exposure/qualified in >1 exposure category and initiation restricted to the date of the drug's latest marketing authorization | -6 982 | 25 704 | -21,36 % |
| Excluded based on missing Age / Gender | 0 | 25 704 | 0,00 % |
| Excluded based on Inclusion - Age >= min Study Age (18y) | -4 | 25 700 | -0,02 % |
| Excluded based on Inclusion - T2D | -1 673 | 24 027 | -6,51 % |
| Excluded based in Inclusion - min 2 registry entries in past year | -386 | 23 641 | -1,61 % |
| Excluded based on Inclusion - high risk of cardiovascular event | -9 373 | 14 268 | -39,65 % |
| Excluded based on exclusion criteria for past 180 days: Obesity, Liver disease, Blood disease, Substance/Alcohol abuse, Pregnancy/Use of Contraceptive | -628 | 13 640 | -4,40 % |
| Excluded based on Exclusion - History of Malignant Neoplasm past 5 years | -1 523 | 12 117 | -11,17 % |
| Excluded based on Exclusion - Chronic Malabsorption | -8 | 12 109 | -0,07 % |
| Excluded based on Exclusion - Anti-obseity drugs | 0 | 12 109 | 0,00 % |
| Excluded based on Exclusion - Acute coronary syndrome, stroke, or transient ischemic attack within 2 months | -262 | 11 847 | -2,16 % |
| Excluded based on Exclusion - Use of corticosteroids | -496 | 11 351 | -4,19 % |
| Excluded based on CCI >= 10 | -2 | 11 349 | -0,02 % |

| | |
|---|---|
| **Final Cohort after incl/excl criteria:** | **11 349** |
| Exposure (Empagliflozin) | 4 630 |
| Comparator (DPP4i) | 6 719 |

| | |
|---|---|
| T2D | Type 2 diabetes |
| CCI | Charlson comorbidity score |
| DPP4i | Dipeptidyl peptidase 4 inhibitor |

**Supplementary Table 5 | Flowchart of the study cohort assembly for the Tecos trial emulation.**
The table lists the included eligibility criteria and the number of the excluded and remaining patients at each step.

| | Excluded patients | Remaining patients | % discarded |
|---|---|---|---|
| All patients | - | 425 483 | |
| Patients who used exposure or a reference | -397 051 | 28 432 | -93,32 % |
| Excluded due to prior use of referent/exposure/qualified in >1 exposure category and initiation restricted to the date of the drug's latest marketing authorization | -7 935 | 20 497 | -27,91 % |
| Excluded based on missing Age / Gender | 0 | 20 497 | 0,00 % |
| Excluded based on Inclusion - Age >= min Study Age (18y) | -2 | 20 495 | -0,01 % |
| Excluded based on Inclusion - T2D | -1 359 | 19 136 | -6,63 % |
| Excluded based in Inclusion - min 2 registry entries in past year | -241 | 18 895 | -1,26 % |
| Excluded based on Inclusion - Rx for Metformin/Insulin/Piogliatzone/SU | -6 685 | 12 210 | -35,38 % |
| Excluded based on Inclusion - Age 50 OR one major CV Risk factor in past 365 days (MI, Revascularization, Coronary Angioplasty (PTCA), Bypass, Ischemic Stroke, Atherosclerosis, Amputation, Stenosis) | -1 484 | 10 726 | -12,15 % |
| Excluded based on Exclusion - T1D, Hypoglycaemia, Ketoacidosis | -9 | 10 717 | -0,08 % |
| Excluded based on Exclusion - Pregnancy/Use of Contraceptive and Liver Cirrhosis | -15 | 10 702 | -0,14 % |
| Excluded based on Exclusion - DPP4i, GLP-1, Thiazolidinediones (other than Pioglitazone) | -226 | 10 476 | -2,11 % |
| Excluded based on CCI >= 10 | -7 | 10 469 | -0,07 % |

| | |
|---|---|
| **Final Cohort after incl/excl criteria:** | **10 469** |
| Exposure (Sitagliptin) | 8 174 |
| Reference (2nd gen Sulfonylurea) | 2 326 |

| | |
|---|---|
| T2D | Type 2 diabetes |
| T1D | Type 1 diabetes |
| Rx | Prescription medicine |
| SU | Sulfonylurea |
| CV | Cardiovacular |
| MI | Myocardial infarction |
| PTCA | Percutaneous transluminal coronary angioplasty |
| CCI | Charlson comorbidity score |
| DPP4i | Dipeptidyl peptidase 4 inhibitor |
| GLP-1 | Glucagon-like peptide 1 |

**Supplementary Table 6 | Flowchart of the study cohort assembly for the Aristotle trial emulation.**
The table lists the included eligibility criteria and the number of the excluded and remaining patients at each step.

| | Excluded patients | Remaining patients | % discarded |
|---|---|---|---|
| All patients | - | 425 483 | |
| Patients who used exposure or a reference | -368 083 | 57 400 | -86,51 % |
| Excluded due to prior use of referent/exposure/qualified in >1 exposure category and initiation restricted to the date of the drug's latest marketing authorization | -16 532 | 40 868 | -28,80 % |
| Excluded based on missing Age / Gender | 0 | 40 868 | 0,00 % |
| Excluded based on Inclusion - Age >= min Study Age (18y) | -75 | 40 793 | -0,18 % |
| Excluded based on Inclusion - AF | -22 776 | 18 017 | -55,83 % |
| Excluded based on Inclusion - min 2 registry entries in past year | -374 | 17 643 | -2,08 % |
| Excluded based on Inclusion - Stroke Risk Factor (Age over 75, MI, TIA, Embolism, HF, DM, Stroke, HTN, HTN meds) | -315 | 17 328 | -1,79 % |
| Excluded based on Exclusion - Ablation, Low platelet /Hemoglobin, Liver disease /insufficiency, Kidney disease, Cancer Alcohol/Substance abuse | -1 522 | 15 806 | -8,78 % |
| Excluded based on Exclusion - Pregnancy/Use of Contraceptive | -5 | 15 801 | -0,03 % |
| Excluded based on Exclusion - Cancer(wide) | -1 179 | 14 622 | -7,46 % |
| Excluded based on CCI >= 10 | -1 | 14 621 | -0,01 % |

| | |
|---|---|
| **Final Cohort after incl/excl criteria:** | **14 621** |
| Exposure (Apixaban) | 8 995 |
| Reference (Warfarin) | 5 626 |

| | |
|---|---|
| AF | Atrial fibrillation |
| MI | Myocardial infarction |
| TIA | Transient ischemic attack |
| HF | Heart failure |
| DM | Diabetes mellitus |
| HTN | Hypertension |
| CCI | Charlson comorbidity score |

**Supplementary Table 7 | Flowchart of the study cohort assembly for the Rocket trial emulation.**
The table lists the included eligibility criteria and the number of the excluded and remaining patients at each step.

| | Excluded patients | Remaining patients | % discarded |
|---|---|---|---|
| All patients | - | 425 483 | |
| Patients who used exposure or a reference | -363 115 | 62 368 | -85,34 % |
| Excluded due to prior use of referent/exposure/qualified in >1 exposure category and initiation restricted to the date of the drug's latest marketing authorization | -13 959 | 48 409 | -22,38 % |
| Excluded based on missing Age / Gender | 0 | 48 409 | 0,00 % |
| Excluded based on Inclusion - Age >= min Study Age (18y) | -85 | 48 324 | -0,18 % |
| Excluded based on Inclusion - AF (measured min 2x in past 365 days) | -32 673 | 15 651 | -67,61 % |
| Excluded based on Inclusion - min 2 registry entries in past year | -400 | 15 251 | -2,56 % |
| Excluded based on Inclusion - Stroke Risk Factor (either one of the following: Stroke, TIA, Embolus OR two of the following: Age >= 75, HF, LVEF ≤35%, DM, HTN) | -8 840 | 6 411 | -57,96 % |
| Excluded based on exclusion criteria for past 180 days: Mitral Stenosis, Prosthetic Heart valve, Thrombus, Endocarditis, Bleeding, PE/DVT, Anemia, Pregnancy/Use of Contraceptive, HIV, CKD, Dialysis, Liver disease, Alcohol/Substance abuse | -622 | 5 789 | -9,70 % |
| Excluded based on Exclusion - Stroke | -806 | 4 983 | -13,92 % |
| Excluded based on exclusion criteria for past 30 days: Haemorrhagic disorder, Intracranial neoplasms, Cerebral aneurysm, Thrombocytopenia | -8 | 4 975 | -0,16 % |
| Excluded based on Exclusion - CYP Inhibitors/Inducers, TIA | -49 | 4926 | -0,98 % |
| Excluded based on CCI >= 10 | -1 | 4 925 | -0,02 % |

| | |
|---|---|
| **Final Cohort after incl/excl criteria:** | **4 925** |
| Exposure (Rivaroxaban) | 1 917 |
| Reference (Warfarin) | 3 008 |

| | |
|---|---|
| AF | Atrial fibrillation |
| MI | Myocardial infarction |
| TIA | Transient ischemic attack |
| HF | Heart failure |
| LVEF | Left ventricular ejection fraction |
| DM | Diabetes mellitus |
| HTN | Hypertension |
| PE | Pulmonary embolism |
| DVT | Deep vein thrombosis |
| HIV | Human immunodeficiency virus |
| CKD | Chronic kidney disease |
| CCI | Charlson comorbidity score |

**Supplementary Table 8 | Propensity score matching statistics for the Empareg trial emulation.**
The prevalence of selected covariates that were included in the propensity score estimation and matching in both trial groups.
Standardized mean difference indicates the balance of the covariate between both trial groups. The standardized mean difference was calculated using the Cohen's d for continuous variables and the Cohen's h for binary variables.

| | Unmatched | | | PS-matched | | |
|---|---|---|---|---|---|---|
| **Covariate** | **Empagliflozin** | **DPP4-inhibitors** | **Std. Diff.** | **Empagliflozin** | **DPP4-inhibitors** | **Std. Diff.** |
| **Number of patients** | 4630 | 6719 | | 2261 | 2261 | |
| **Female gender** | 38.70% | 40.20% | -0.031 | 39.80% | 40.20% | -0.01 |
| **Mean Age (SD)** | 65.86 (10.47) | 71.48 (10.59) | -0.534 | 66.95 (10.87) | 67.63 (11.03) | -0.063 |
| **Mean charles comorbidity index (SD)** | 0.83 (1.08) | 0.83 (1.23) | 0.005 | 0.85 (1.11) | 0.85 (1.17) | -0.001 |
| **Mean birthyear (SD)** | 1953.47 (10.27) | 1944.88 (10.7) | 0.819 | 1951.72 (10.69) | 1950.9 (10.4) | 0.077 |
| **Combinations of oral blood glucose lowering drugs** | 16.20% | 6.20% | 0.327 | 15.70% | 13.60% | 0.059 |
| **Antihypertensive medication** | 98.3% | 97.9% | 0.027 | 98.10% | 98.30% | -0.01 |
| **Benzodiazepine use** | 5.6% | 6.5% | -0.041 | 5.80% | 5.50% | 0.013 |
| **Statin medication** | 74.7% | 72.6% | 0.048 | 74.00% | 72.90% | 0.026 |
| **Biguanides** | 74.3% | 67.8% | 0.144 | 73.90% | 75.20% | -0.029 |
| **Insulins** | 24.9% | 23.9% | 0.022 | 24.90% | 23.30% | 0.038 |
| **Opioids** | 17.5% | 17.1% | 0.01 | 17.90% | 18.00% | -0.001 |
| **NSAIDs** | 29.8% | 20.9% | 0.205 | 28.00% | 27.10% | 0.019 |
| **Diabetic nephropathy** | 0.8% | 1.7% | -0.085 | 1.00% | 0.90% | 0.005 |
| **Chronic obstructive pulmonary disease** | 1.7% | 2.9% | -0.08 | 1.80% | 2.00% | -0.016 |
| **Other diseases of the respiratory system** | 9.7% | 11.4% | -0.054 | 9.80% | 10.70% | -0.032 |
| **Cardiovascular event** | 5.4% | 3.0% | 0.12 | 4.40% | 3.90% | 0.024 |
| **Cardiovascular diseases** | 6.8% | 5.1% | 0.072 | 6.10% | 5.40% | 0.029 |
| **Sulfonylureas** | 2.4% | 5.8% | -0.172 | 2.70% | 3.10% | -0.026 |
| **Other glucose lowering medication** | 2.6% | 2.8% | -0.009 | 2.90% | 4.70% | -0.093 |
| **Asthma** | 6.6% | 5.3% | 0.055 | 6.30% | 6.30% | 0 |
| **Chronic kidney disease** | 0.6% | 3.7% | -0.23 | 1.00% | 1.20% | -0.017 |
| **Diabetic neuropathy** | 0.5% | 0.5% | -0.007 | 0.40% | 0.40% | 0.007 |
| **Strong smoking dependency** | 0.1% | 0.1% | 0.004 | 0.00% | 0.10% | -0.031 |
| **Hypoglycaemia** | 0.0% | 0.1% | -0.025 | 0.00% | 0.00% | 0 |
| **Neovascular glaucoma** | 0.0% | 0.1% | -0.044 | 0.00% | 0.10% | -0.017 |
| **Gangrene** | 0.0% | 0.0% | -0.035 | 0.00% | 0.00% | 0 |
| **Antithrombotic medication** | 34.8% | 36.5% | -0.036 | 34.10% | 34.10% | 0 |

| | |
|---|---|
| SD | Standard deviation |
| Std. Diff. | Standardized mean difference |
| PS | Propensity score |
| DPP4 | Dipeptidyl peptidase 4 |
| NSAIDs | nonsteroidal anti-inflammatory drugs |

**Supplementary Table 9 | Propensity score matching statistics for the Tecos trial emulation.**
The prevalence of selected covariates that were included in the propensity score estimation and matching in both trial groups.
Standardized mean difference indicates the balance of the covariate between both trial groups. The standardized mean difference was calculated using the Cohen's d for continuous variables and the Cohen's h for binary variables.

| | Unmatched | | | PS-matched | | |
|---|---|---|---|---|---|---|
| **Covariate** | **Sitagliptin** | **2G Sulfonylureas** | **Std. Diff.** | **Sitagliptin** | **2G Sulfonylureas** | **Std. Diff.** |
| **Number of patients** | 8143 | 2326 | | 547 | 547 | |
| **Female gender** | 45.3% | 33.5% | 0.243 | 41.5% | 42.0% | -0.011 |
| **Mean Age (SD)** | 66.02 (8.89) | 68.19 (9.4) | -0.237 | 65.55 (9) | 64.29 (8.27) | 0.146 |
| **Mean charles comorbidity index (SD)** | 0.62 (1.07) | 0.38 (0.84) | 0.248 | 0.46 (0.88) | 0.42 (0.93) | 0.04 |
| **Mean birthyear (SD)** | 1947.9 (9.16) | 1939.37 (10.03) | 0.888 | 1946.3 (9.64) | 1946.11 (9.01) | 0.02 |
| **Biguanides** | 91.9% | 78.3% | 0.391 | 90.7% | 94.1% | -0.132 |
| **Antihypertensive medication** | 84.3% | 84.2% | 0.003 | 86.1% | 84.5% | 0.046 |
| **NSAIDs** | 32.0% | 30.9% | 0.024 | 34.6% | 35.3% | -0.015 |
| **Statin medication** | 66.7% | 61.7% | 0.106 | 66.7% | 66.2% | 0.012 |
| **Insulins** | 18.9% | 23.8% | -0.119 | 13.2% | 13.2% | 0 |
| **Benzodiazepine use** | 6.5% | 6.8% | -0.013 | 5.5% | 7.1% | -0.068 |
| **Opioids** | 15.0% | 6.7% | 0.271 | 10.2% | 12.4% | -0.069 |
| **Other glucose lowering medication** | 2.0% | 1.3% | 0.053 | 2.6% | 3.3% | -0.043 |
| **Chronic obstructive pulmonary disease** | 1.8% | 3.0% | -0.078 | 2.6% | 2.7% | -0.011 |
| **Other diseases of the respiratory system** | 8.5% | 7.8% | 0.023 | 6.8% | 8.4% | -0.062 |
| **Cardiovascular event** | 1.7% | 3.3% | -0.105 | 2.0% | 2.4% | -0.025 |
| **Cardiovascular diseases** | 3.2% | 5.1% | -0.098 | 3.7% | 2.9% | 0.041 |
| **Asthma** | 4.8% | 2.6% | 0.117 | 3.3% | 3.8% | -0.03 |
| **Combinations of oral blood glucose lowering drugs** | 3.2% | 3.8% | -0.035 | 5.5% | 6.8% | -0.053 |
| **Chronic kidney disease** | 0.6% | 0.3% | 0.046 | 0.7% | 0.5% | 0.023 |
| **Diabetic nephropathy** | 0.7% | 0.7% | -0.003 | 0.2% | 0.2% | 0 |
| **Neovascular glaucoma** | 0.1% | 0.2% | -0.034 | 0.2% | 0.2% | 0 |
| **Diabetic neuropathy** | 0.4% | 0.3% | 0.023 | 0.7% | 0.4% | 0.05 |
| **SGLT2-inhibitors** | 0.1% | 0.0% | 0.035 | 0.0% | 0.2% | -0.086 |
| **Strong smoking dependency** | 0.0% | 0.1% | -0.014 | 0.0% | 0.0% | 0 |
| **Antithrombotic medication** | 21.9% | 23.2% | -0.031 | 18.3% | 18.3% | 0 |

| | |
|---|---|
| SD | Standard deviation |
| Std. Diff. | Standardized mean difference |
| PS | Propensity score |
| 2G | Second generation |
| NSAIDs | nonsteroidal anti-inflammatory drugs |
| SGLT2 | Sodium-glucose cotransporter-2 |

**Supplementary Table 10 | Propensity score matching statistics for the Aristotle trial emulation.**
The prevalence of selected covariates that were included in the propensity score estimation and matching in both trial groups.
Standardized mean difference indicates the balance of the covariate between both trial groups. The standardized mean difference was calculated using the Cohen's d for continuous variables and the Cohen's h for binary variables.

| | Unmatched | | | PS-matched | | |
|---|---|---|---|---|---|---|
| **Covariate** | **Apixaban** | **Warfarin** | **Std. Diff.** | **Apixaban** | **Warfarin** | **Std. Diff.** |
| **Number of patients** | 8995 | 5626 | | 1146 | 1146 | |
| **Female gender** | 46.5% | 39.2% | 0.148 | 42.9% | 42.7% | 0.005 |
| **Mean Age (SD)** | 74.59 (9.89) | 70.77 (10.27) | 0.378 | 72.27 (10.45) | 72.47 (10.19) | -0.02 |
| **Mean charles comorbidity index (SD)** | 0.8 (1.1) | 0.81 (1.15) | -0.005 | 0.83 (1.12) | 0.84 (1.13) | -0.009 |
| **Mean birthyear (SD)** | 1944.84 (9.9) | 1942.73 (10.4) | 0.208 | 1944.77 (10.42) | 1944.48 (10.67) | 0.028 |
| **Diabetes mellitus** | 25.9% | 23.1% | 0.065 | 22.7% | 23.2% | -0.012 |
| **Asthma** | 7.2% | 6.7% | 0.02 | 7.4% | 7.1% | 0.013 |
| **Chronic obstructive pulmonary disease** | 3.4% | 4.4% | -0.053 | 4.1% | 3.8% | 0.013 |
| **Other diseases of the respiratory system** | 16.8% | 18.5% | -0.044 | 19.1% | 19.3% | -0.004 |
| **Antihypertensive medication** | 96.1% | 96.4% | -0.018 | 95.8% | 96.1% | -0.013 |
| **Statin medication** | 58.7% | 53.0% | 0.116 | 54.2% | 53.1% | 0.023 |
| **Transient ischemic attack** | 3.8% | 3.9% | -0.001 | 4.0% | 4.7% | -0.034 |
| **NSAIDs** | 23.3% | 23.2% | 0.002 | 25.2% | 25.9% | -0.016 |
| **Heart failure** | 30.9% | 31.4% | -0.009 | 31.2% | 31.5% | -0.006 |
| **Benzodiazepine use** | 6.1% | 6.2% | -0.004 | 6.4% | 6.1% | 0.011 |
| **Diabetes medication** | 25.0% | 22.6% | 0.059 | 22.2% | 22.8% | -0.015 |
| **Opioids** | 16.6% | 17.3% | -0.017 | 15.7% | 16.3% | -0.017 |
| **Cardiovascular event** | 6.7% | 6.3% | 0.017 | 6.4% | 6.7% | -0.014 |
| **Cardiovascular diseases** | 14.2% | 12.1% | 0.061 | 15.3% | 14.0% | 0.035 |
| **SGLT2-inhibitors** | 8.1% | 5.2% | 0.115 | 6.6% | 7.1% | -0.017 |
| **Diabetic nephropathy** | 0.4% | 0.6% | -0.026 | 0.6% | 0.7% | -0.011 |
| **Chronic kidney disease** | 2.3% | 2.4% | -0.008 | 2.9% | 3.2% | -0.02 |
| **Dronedarone medication** | 0.5% | 1.1% | -0.064 | 0.8% | 0.3% | 0.059 |
| **Strong smoking dependency** | 0.0% | 0.0% | -0.016 | 0.0% | 0.0% | 0 |
| **Diabetic neuropathy** | 0.1% | 0.3% | -0.04 | 0.1% | 0.2% | -0.024 |
| **Hypoglycaemia** | 0.1% | 0.1% | 0.002 | 0.0% | 0.1% | -0.059 |
| **Neovascular glaucoma** | 0.2% | 0.1% | 0.011 | 0.3% | 0.3% | 0.016 |

| | |
|---|---|
| SD | Standard deviation |
| Std. Diff. | Standardized mean difference |
| PS | Propensity score |
| NSAIDs | nonsteroidal anti-inflammatory drugs |
| SGLT2 | Sodium-glucose cotransporter-2 |

**Supplementary Table 11 | Propensity score matching statistics for the Rocket trial emulation.**
The prevalence of selected covariates that were included in the propensity score estimation and matching in both trial groups.
Standardized mean difference indicates the balance of the covariate between both trial groups. The standardized mean difference was calculated using the Cohen's d for continuous variables and the Cohen's h for binary variables.

| | Unmatched | | | PS-matched | | |
|---|---|---|---|---|---|---|
| **Covariate** | **Rivaroxaban** | **Warfarin** | **Std. Diff.** | **Rivaroxaban** | **Warfarin** | **Std. Diff.** |
| **Number of patients** | 1917 | 3008 | | 515 | 515 | |
| **Female gender** | 43.7% | 40.6% | 0.064 | 41.0% | 41.9% | -0.02 |
| **Mean Age (SD)** | 76.9 (8.6) | 76.04 (8.75) | 0.099 | 76.66 (9.01) | 77.16 (8.83) | -0.056 |
| **Mean charles comorbidity index (SD)** | 1.1 (1.36) | 1.11 (1.28) | -0.006 | 1.15 (1.29) | 1.15 (1.24) | -0.003 |
| **Mean birthyear (SD)** | 1941.45 (8.63) | 1936.47 (8.72) | 0.575 | 1939.82 (9.16) | 1939.05 (8.77) | 0.087 |
| **Heart failure** | 40.4% | 51.5% | -0.223 | 43.3% | 46.8% | -0.07 |
| **Antihypertensive medication** | 96.5% | 97.3% | -0.048 | 96.7% | 97.7% | -0.059 |
| **Opioids** | 19.8% | 19.8% | -0.001 | 19.6% | 20.8% | -0.029 |
| **Diabetes medication** | 38.5% | 33.7% | 0.1 | 34.6% | 34.8% | -0.004 |
| **Diabetes mellitus** | 39.9% | 34.3% | 0.115 | 36.1% | 35.7% | 0.008 |
| **Cardiovascular event** | 4.5% | 8.3% | -0.156 | 6.2% | 5.8% | 0.016 |
| **Cardiovascular diseases** | 8.6% | 12.8% | -0.138 | 9.5% | 9.1% | 0.013 |
| **Statin medication** | 60.2% | 62.7% | -0.052 | 61.0% | 59.8% | 0.024 |
| **Benzodiazepine use** | 6.1% | 7.1% | -0.042 | 5.8% | 6.0% | -0.008 |
| **NSAIDs** | 20.8% | 18.2% | 0.066 | 22.5% | 21.7% | 0.019 |
| **SGLT2-inhibitors** | 11.5% | 6.4% | 0.178 | 9.3% | 9.9% | -0.02 |
| **Asthma** | 7.3% | 7.7% | -0.018 | 9.7% | 8.5% | 0.04 |
| **Other diseases of the respiratory system** | 20.1% | 23.3% | -0.078 | 23.7% | 23.5% | 0.005 |
| **Chronic obstructive pulmonary disease** | 4.2% | 6.6% | -0.105 | 6.2% | 5.8% | 0.016 |
| **Transient ischemic attack** | 2.0% | 3.2% | -0.079 | 2.5% | 1.2% | 0.103 |
| **Diabetic nephropathy** | 0.7% | 1.3% | -0.066 | 1.2% | 1.4% | -0.017 |
| **Dronedarone medication** | 0.1% | 0.9% | -0.141 | 0.2% | 0.0% | 0.088 |
| **Chronic kidney disease** | 0.4% | 0.2% | 0.032 | 0.6% | 0.4% | 0.028 |
| **Diabetic neuropathy** | 0.2% | 0.4% | -0.052 | 0.2% | 0.4% | -0.037 |
| **Neovascular glaucoma** | 0.1% | 0.2% | -0.017 | 0.0% | 0.0% | 0 |
| **Hypoglycaemia** | 0.1% | 0.2% | -0.044 | 0.2% | 0.2% | 0 |
| **Strong smoking dependency** | 0.0% | 0.0% | -0.036 | 0.0% | 0.0% | 0 |
| **Diuretics** | 38.0% | 47.4% | -0.19 | 40.4% | 43.1% | -0.055 |
| **Platelet aggregation inhibitors** | 16.4% | 14.8% | 0.044 | 16.1% | 17.1% | -0.026 |
| **Corticosteroids for systemic use** | 15.9% | 16.0% | -0.003 | 17.5% | 18.4% | -0.025 |

| | |
|---|---|
| SD | Standard deviation |
| Std. Diff. | Standardized mean difference |
| PS | Propensity score |
| NSAIDs | nonsteroidal anti-inflammatory drugs |
| SGLT2 | Sodium-glucose cotransporter-2 |

**Supplementary Table 12 | Genome-wide association study summary statistics used to compute polygenic risk scores**

| Phenotype | Link to paper/reference | 1st author and year | GWAS Trait | Controls | Cases | Link to sumstats | Comment |
|---|---|---|---|---|---|---|---|
| Atrial fibrillation | https://doi.org/10.1038/s41588-01 | Roselli et al. 2018 | Atrial fibrillation | 522,744 | 65,446 | http://ftp.ebi.ac.uk/pub/d | - |
| Asthma | https://doi.org/10.1038/s41467-020-1 | Han et al. 2020 | Asthma | 447,859 | 88,486 | http://ftp.ebi.ac.uk/pub/d | - |
| ALT enzyme | https://pan.ukbb.broadinstitute.org | Pan-UK Biobank | Alanine aminotransferase | 400,822 | | https://docs.google.co | only European subset |
| AST enzyme | https://pan.ukbb.broadinstitute.org | Pan-UK Biobank | Aspartate aminotransferase | 399482 | | https://docs.google.co | only European subset |
| Body mass index | https://doi.org/10.1038/nature141 | Locke et al. 2015 | Body mass index | 236,781 | | https://ftp.ebi.ac.uk/pul | |
| C-reactive protein | https://pan.ukbb.broadinstitute.org | Pan-UK Biobank | C-reactive protein | 400,094 | | https://docs.google.co | only European subset |
| Chronic kidney disease | https://doi.org/10.1038/s41588-01 | Wuttke et al. 2019 | Chronic kidney disease | 561,055 | 64,164 | https://ckdgen.imbi.uni | - |
| Coronary heart disease | https://www.nature.com/articles/ng.3 | Nelson et al. 2017 | Coronary artery disease | 260,875 | 71,602 | http://www.cardiograml | - |
| Education | https://doi.org/10.1038/s41588-01 | Lee et al. 2018 | Educational attainment | 1,131,881 | | http://www.thessgac.o | - |
| Heart failure | https://doi.org/10.1038/s41467-01 | Shah et al. 2020 | Heart failure | 930,014 | 47,309 | https://cvd.hugeamp.o | |
| HbA1c | https://pan.ukbb.broadinstitute.org | Pan-UK Biobank | Glycated haemoglobin (HbA1c) | 400,825 | | https://docs.google.co | only European subset |
| HDL | https://pan.ukbb.broadinstitute.org | Pan-UK Biobank | HDL cholesterol | 367,021 | | https://docs.google.co | only European subset |
| LDL | https://pan.ukbb.broadinstitute.org | Pan-UK Biobank | LDL direct | 400,223 | | https://docs.google.co | only European subset |
| Liver disease | https://journals.lww.com/hepcomn | Fairfield et al. 2021 | Nonalcoholic fatty liver disease (NAFLD | 373,227 | 4,761 | - | |
| Major depression | https://doi.org/10.1038/s41588-018-0 | Wray et al 2018 | Major depressive disorder | 113,154 | 59,851 | https://figshare.com/artic | - |
| Subararchnoid hemorrhage | https://doi.org/10.1038/s41588-02 | Bakker et al. 2020 | Subararchnoid hemorrhage | ? | ? | http://www.cerebrovas | |
| Systolic blood pressure | https://pan.ukbb.broadinstitute.org | Pan-UK Biobank | Systolic blood pressure | 396,663 | | https://docs.google.co | only European subset |
| Stroke | https://doi.org/10.1038/s41588-01 | Malik et al. 2018 | All strokes | 454,450 | 67,162 | https://ftp.ebi.ac.uk/pul | |
| Type 2 diabtetes | https://doi.org/10.1038/s41586-02 | Suzuki et al. 2024 | Type 2 diabetes | 2,107,149 | 428,452 | http://www.diagram-co | |
| Triglycerides | https://pan.ukbb.broadinstitute.org | Pan-UK Biobank | Triglycerides | 400,639 | | https://docs.google.co | only European subset |
