## Supplementary Material for "Integrating genetic data in target trial emulations improves their design and informs the value of polygenic scores for prognostic and predictive enrichment"

### Supplementary Figure 1 – Trial emulation design

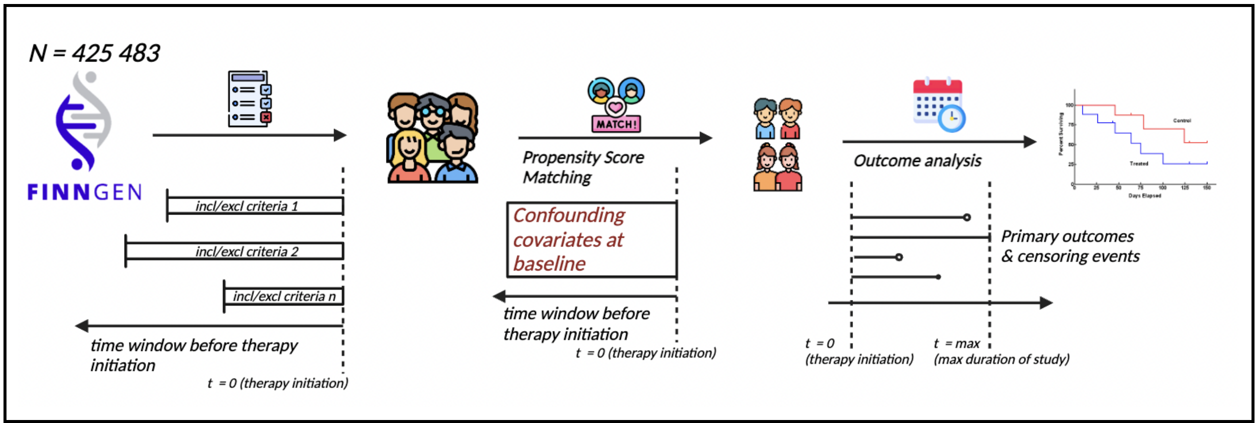


**Supplementary Figure 1. Trial emulation design**
This flowchart depicts the involved steps in closely replicating original randomized controlled trials (RCT) in observational data. Using FinnGen as our data source, we defined the treatment strategies, translated and applied the RCT's eligibility criteria, selected confounding variables and incorporated 1:1 nearest-neighbor propensity score-matching, and performed an outcome analysis after specifying primary endpoints and censoring events, finally obtaining hazard ratios and 95% confidence intervals.
Incl, inclusion; excl, exclusion; t, time.

### Supplementary Figure 2 – Standardized mean difference of 20 polygenic risk scores across different stages of the Tecos trial emulation

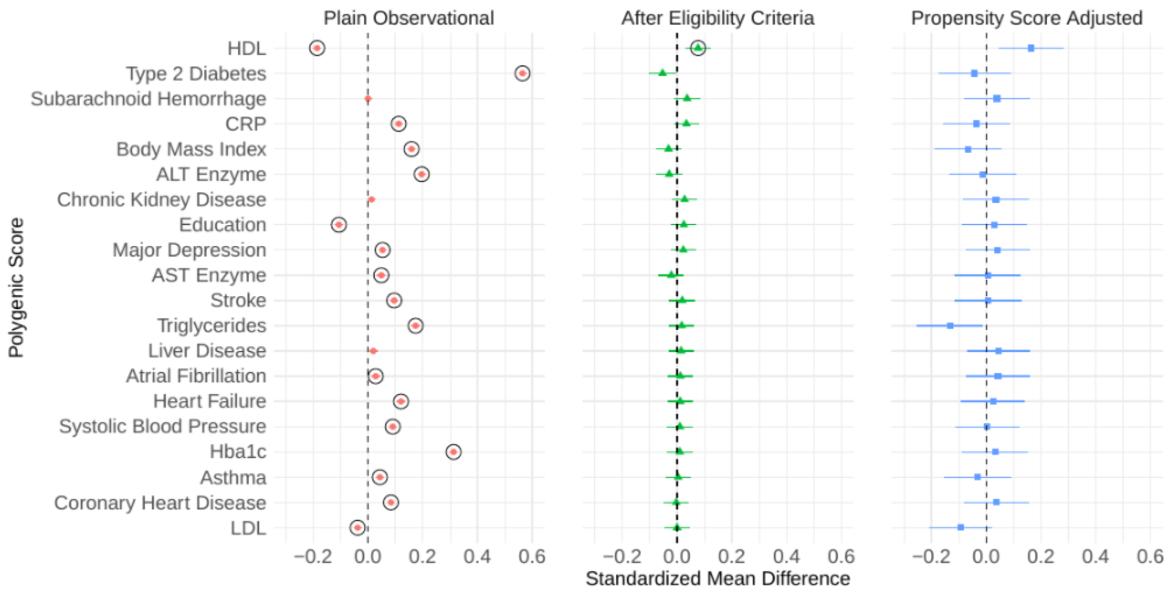


**Supplementary Figure 2. Standardized mean difference of 20 polygenic risk scores across different stages of the Tecos trial emulation**
Plain Observational: initiators of the treatment vs non-initiators; After Eligibility Criteria: Tecos trial emulation cohort after applying inclusion/exclusion criteria and including an active comparator group (2nd generation Sulfonylureas user); Propensity Score Adjusted: Tecos trial emulation cohorts after with inclusion/exclusion criteria and a 1:1 propensity score nearest-neighbor matching for 26 covariates.
The standardized differences in means of the two trial arms are plotted as point estimates and lines representing their 95% CI. A circle around the point estimates represents statistical significance after a Bonferroni-corrected P value threshold ($2.5 \times 10^{-3}$).
ALT, alanine transaminase; AST, aspartate transaminase; Hba1c, glycated hemoglobin A1c; CRP, C-reactive protein; LDL, low-density lipoprotein; HDL, high-density lipoprotein; CI, confidence interval.

### Supplementary Figure 3 – Standardized mean difference of 20 polygenic risk scores across different stages of the Aristotle trial emulation

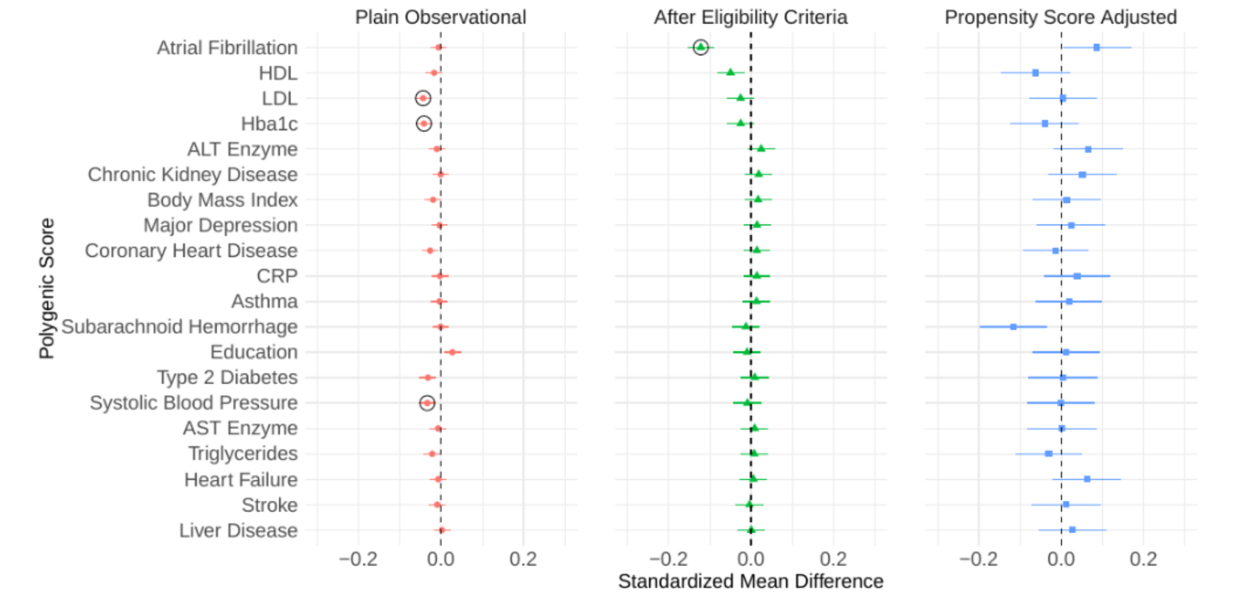


**Supplementary Figure 3. Standardized mean difference of 20 polygenic risk scores across different stages of the Aristotle trial emulation**

Plain Observational: initiators of the treatment (Apixaban) vs initiators of the comparator treatment (Warfarin), reflecting the two trial arms of the original RCT; After Eligibility Criteria: Aristotle trial emulation cohort after applying inclusion/exclusion criteria; Propensity Score Adjusted: Aristotle trial emulation cohorts after with inclusion/exclusion criteria and a 1:1 propensity score nearest-neighbor matching for 26 covariates.

The standardized differences in means of the two trial arms are plotted as point estimates and lines representing their 95% CI. A circle around the point estimates represents statistical significance after a Bonferroni-corrected P value threshold ($2.5 \times 10^{-3}$).

ALT, alanine transaminase; AST, aspartate transaminase; Hba1c, glycated hemoglobin A1c; CRP, C-reactive protein; LDL, low-density lipoprotein; HDL, high-density lipoprotein; RCT, Randomized controlled trial; CI, confidence interval.

### Supplementary Figure 4 – Standardized mean difference of 20 polygenic risk scores across different stages of the Rocket trial emulation

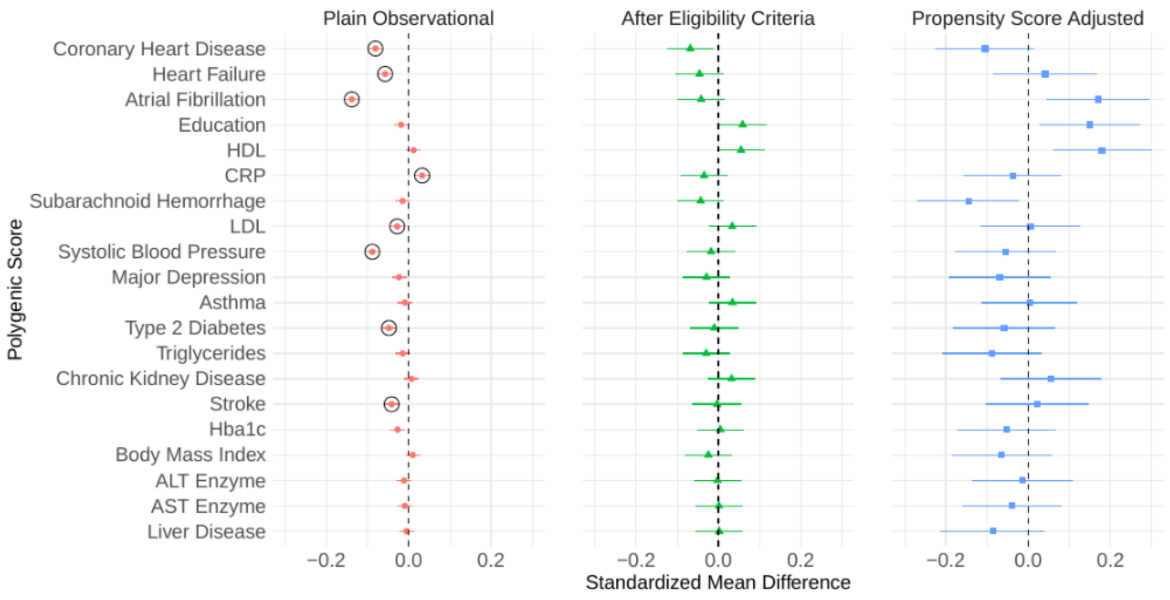


**Supplementary Figure 4. Standardized mean difference of 20 polygenic risk scores across different stages of the Rocket trial emulation**

Plain Observational: initiators of the treatment (Rivaroxaban) vs initiators of the comparator treatment (Warfarin), reflecting the two trial arms of the original RCT; After Eligibility Criteria: Rocket trial emulation cohort after applying inclusion/exclusion criteria; Propensity Score Adjusted: Rocket trial emulation cohorts after with inclusion/exclusion criteria and a 1:1 propensity score nearest-neighbor matching for 30 covariates.

The standardized differences in means of the two trial arms are plotted as point estimates and lines representing their 95% CI. A circle around the point estimates represents statistical significance after a Bonferroni-corrected P value threshold ($2.5 \times 10^{-3}$).

ALT, alanine transaminase; AST, aspartate transaminase; Hba1c, glycated hemoglobin A1c; CRP, C-reactive protein; LDL, low-density lipoprotein; HDL, high-density lipoprotein; RCT, Randomized controlled trial; CI, confidence interval.

### Supplementary Figure 5 – Polygenic Scores induce bias in the association between treatment and outcome

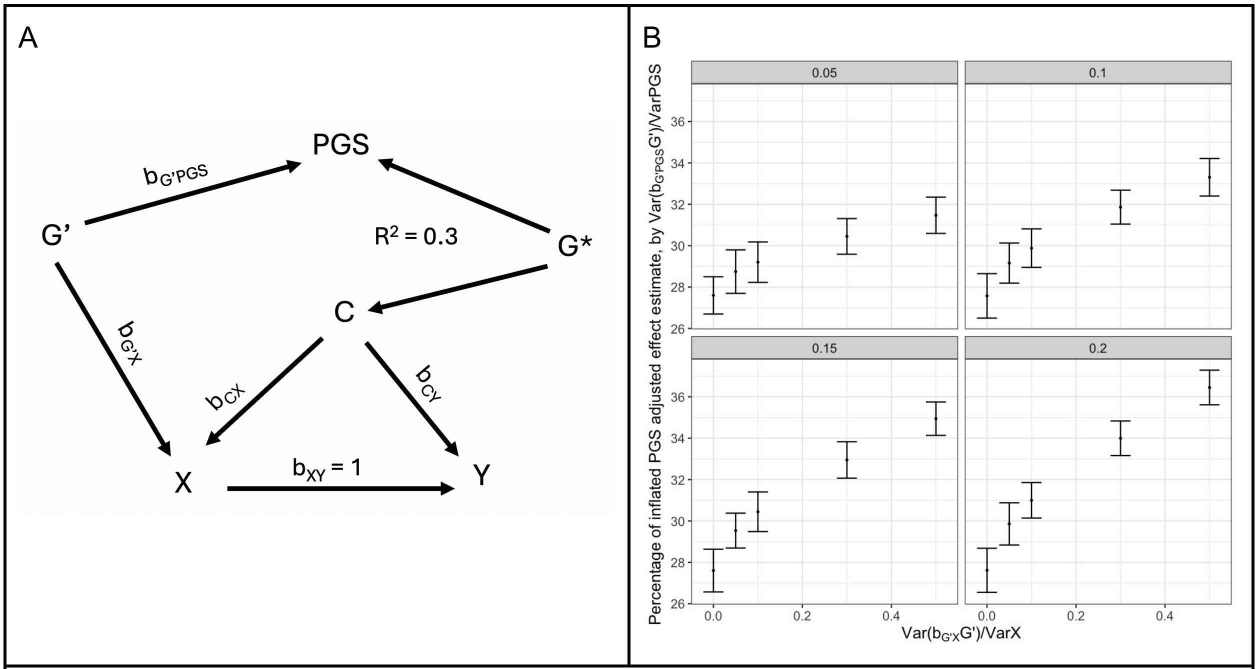


**Supplementary Figure 5. Polygenic Scores induce bias in the association between treatment and outcome.**

A. A directed acyclic graph (DAG) illustrates the embedding of the polygenic score (PGS) in the treatment, outcome, and confounder causal structure. The PGS consists of a genetic component (G*) that is shared with the confounder (C). The effect of G* on the PGS is denoted as $b_{G^*PGS}$ and $b_{G^*C}$ which equals to r/$b_{G^*PGS}$ describes the effect of G* on C, so that the correlation coefficient between PGS and C is r. $r^2$ is fixed as 0.3 in the experiment. The PGS also captures genetic effects (G') that are not causal to the confounder through $b_{G'PGS}$. G' may have a direct effect on the exposure (X) and outcome (Y). We focus on the case where G' only affects X but not Y, and denotes the effect as $b_{G'X}$.The effect of C on X and Y are denoted as $b_{cx}$ and $b_{cy}$ respectively.

B. Simulation study:

We fixed $b_{G^*PGS}^2 = 0.8$ and $r^2 = 0.3$. For simplicity, we only consider the impact of $b_{G'X}$ and $b_{G'PGS}$. Contribution of C on the variance of X and Y are both fixed as 0.3. The true unconfounded effect of X on Y is $b_{xy} = 1$. As a proof of concept, we focus on the scenario where G' negatively impact X, ie. $b_{G'X} < 0$. We investigate changes in estimate bias under various levels of contribution of G' on X, and G' on PGS.

X, treatment; Y, outcome; C, confounder; PGS, polygenic score; r, correlation between PGS and C; $b_{cx}$, effect of C on X ; $b_{cy}$, effect of C on Y, $b_{xy}$, effect of X on Y; $b_{G^*PGS}$, effect of G* on PGS; $b_{G^*C}$, effect of G* on C; $b_{G'PGS}$, effect of G' on PGS; $b_{G'X}$, effect of G' on X; $b_{G'Y}$, effect of G' on Y.

### Supplementary Figure 6 – Evaluating the utility of polygenic scores for confounder adjustment II

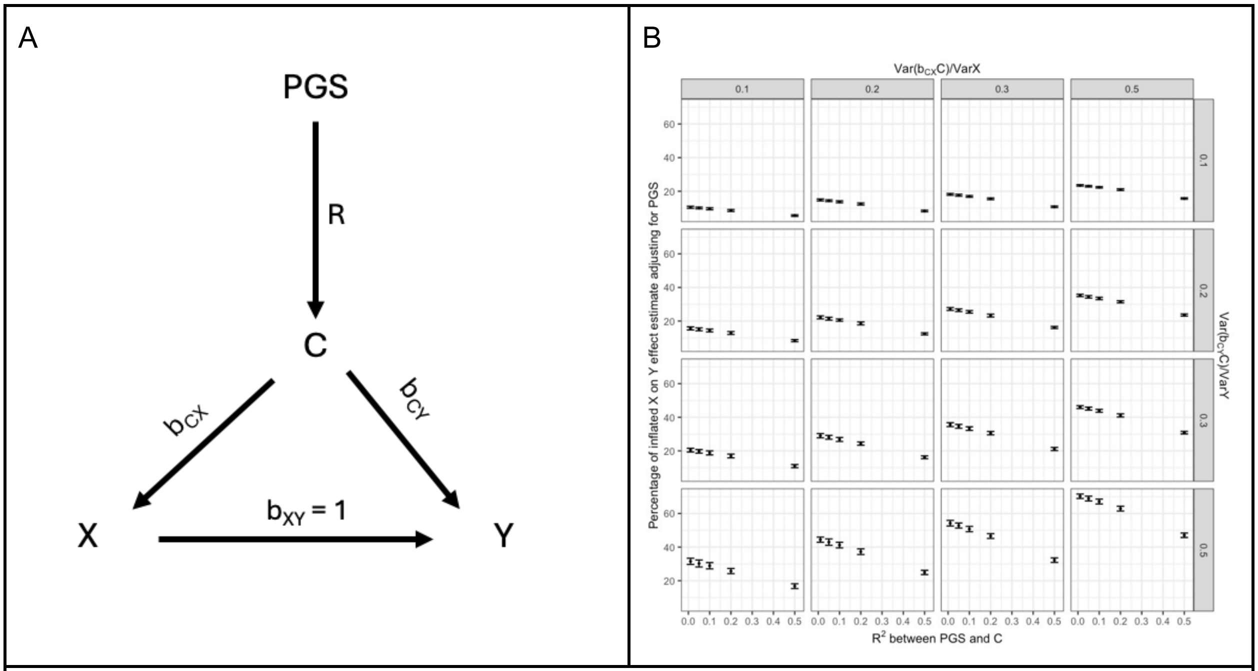


**Supplementary Figure 6. Evaluating the utility of polygenic scores for confounder adjustment II.**
A. A directed acyclic graph (DAG) illustrates the embedding of the polygenic score (PGS) in the treatment, outcome, confounder causal structure. The PGS serves as an imperfect proxy variable for the confounder (C). The effect of the C on the exposure (X) and outcome (Y) are denoted as $b_{cx}$ and $b_{cy}$ , respectively. The true unconfounded effect of X on Y is $b_{xy} = 1$.
B. Simulation study: Under this model (see **Methods**), we changed the correlation between C and PGS simply by varying *R,* and the effect of confounding factor C on X and Y by varying $b_{cx}$ and $b_{cy}$. Under each condition, we measured the observed effect of X on Y, conditioned on PGS, which was an imperfect proxy of C, through linear regression *lm(Y ~ X + PGS)*. We denoted estimate bias as the observed regression coefficient – 1, which is the expected underlying effect of X on Y. The figure shows the estimate bias in respect of the changes in $R^2$, $\frac{bcx2}{Var\ (X)}$ and $\frac{bcy2}{Var\ (X)}$.
X, treatment; Y, outcome; C, confounder; PGS, polygenic score; R, correlation between PGS and C; $b_{cx}$ , effect of C on X ; $b_{cy}$ , effect of C on Y, $b_{xy}$ , effect of X on Y.

### Supplementary Figure 7 – Using mendelian randomization within Empareg trial emulation to identify confounders (adjusted for a C → Y → X effect)

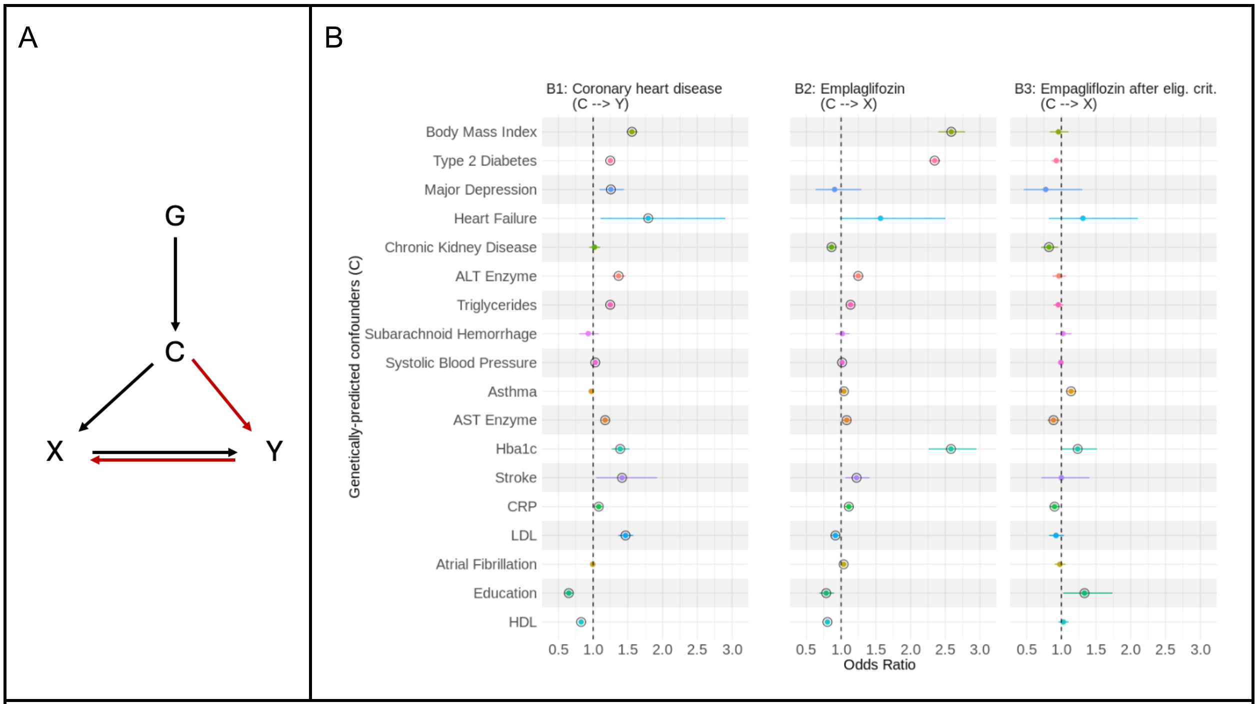


**Supplementary Figure 7. Using mendelian randomization within Empareg trial emulation to identify confounders (adjusted for a C → Y → X effect).**

A. A direct acyclic graph (DAG) illustrating the relationship between treatment initiation (X) and trial outcome (Y), as well as the effect of a genetic instrument (G) of a confounding variable (C) on both the treatment initiation and trial outcome only through the confounding variable. Additionally, a reverse effect of Y on X exists.

B. Results of a mendelian randomization (MR) analysis using inverse variance-weighted to study the causal effects of 18 traits on coronary heart disease, representing the trial outcome, and empagliflozin, representing the treatment initiation. B1. MR for association between 18 traits on coronary artery disease using two-sample MR. B2. MR for association between 18 traits on empagliflozin initiation in the full study population. The effect is adjusted by accounting for the effect of C → Y → X. B3. MR for association between 18 traits on empagliflozin initiation after applying the randomized controlled trial's eligibility criteria. The effect is adjusted by accounting for the effect of C → Y → X.

The point estimates represent the odds ratios with lines representing their 95% confidence interval. For continuous confounders, the odds ratio reflects the change in the outcome variable associated with a 1 SD increase in the exposure variable; for binary confounders, the odds ratio represents the change in the outcome variable when comparing the presence versus the absence of the binary exposure. A circle around the point estimates represents statistical significance with a P value threshold of 5 x 10-2.

ALT, alanine transaminase; AST, aspartate transaminase; Hba1c, glycated hemoglobin A1c; CRP, C-reactive protein; LDL, low-density lipoprotein; HDL, high-density lipoprotein; elig. crit., eligibility criteria; X, treatment; Y, outcome; C, confounder; G, genetic instrument.

### Supplementary Figure 8 – Cohort composition flowchart for Empareg trial emulation

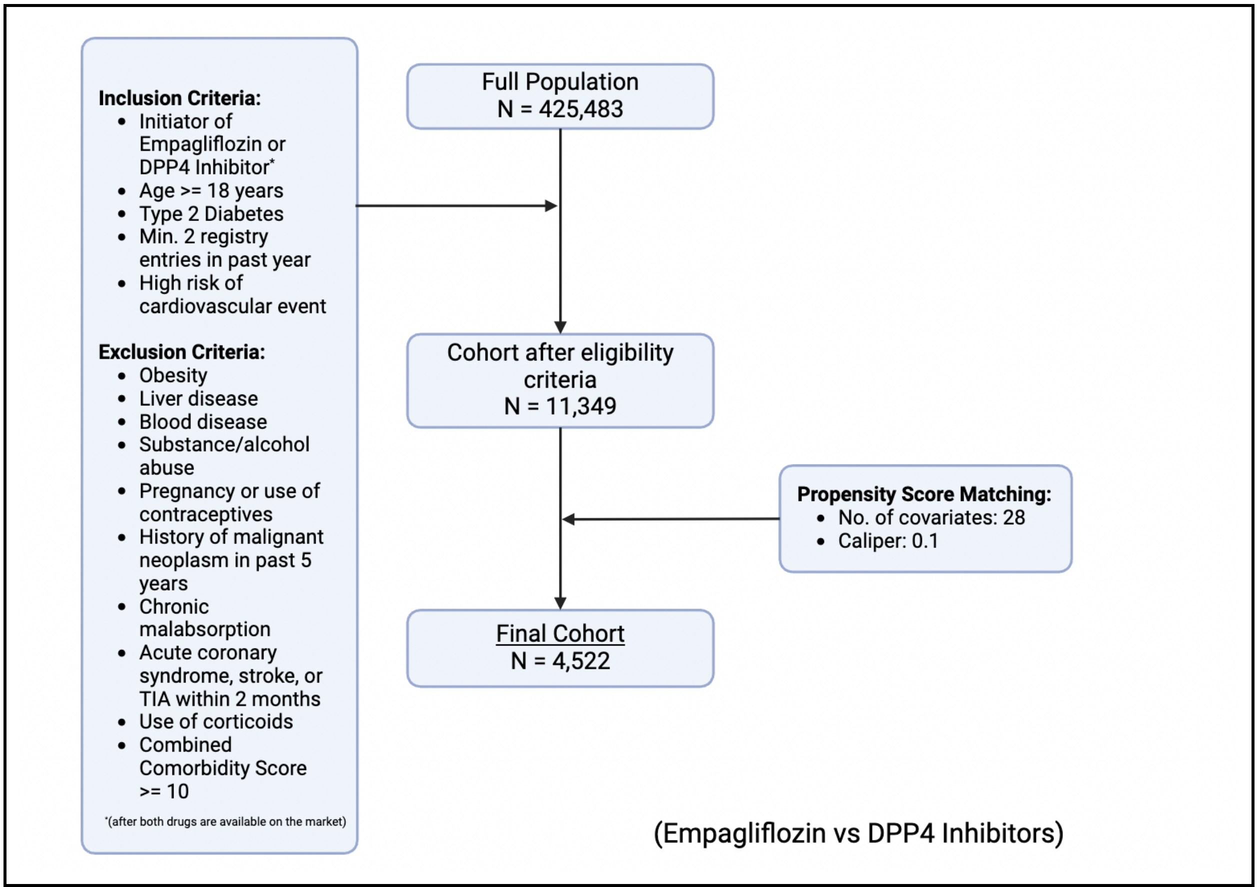


**Supplementary Figure 8. Cohort composition flowchart for Empareg trial emulation**
This flowchart illustrates the sample size changes along the incorporation of the eligibility (inclusion/exclusion) criteria and propensity score matching during the trial emulation.
A flowchart of the cohort formation can be found in **Supplementary Table 4**.
Details on the covariates used for the propensity score matching can be found in **Supplementary Table 8**.

### Supplementary Figure 9 – Cohort composition flowchart for Tecos trial emulation

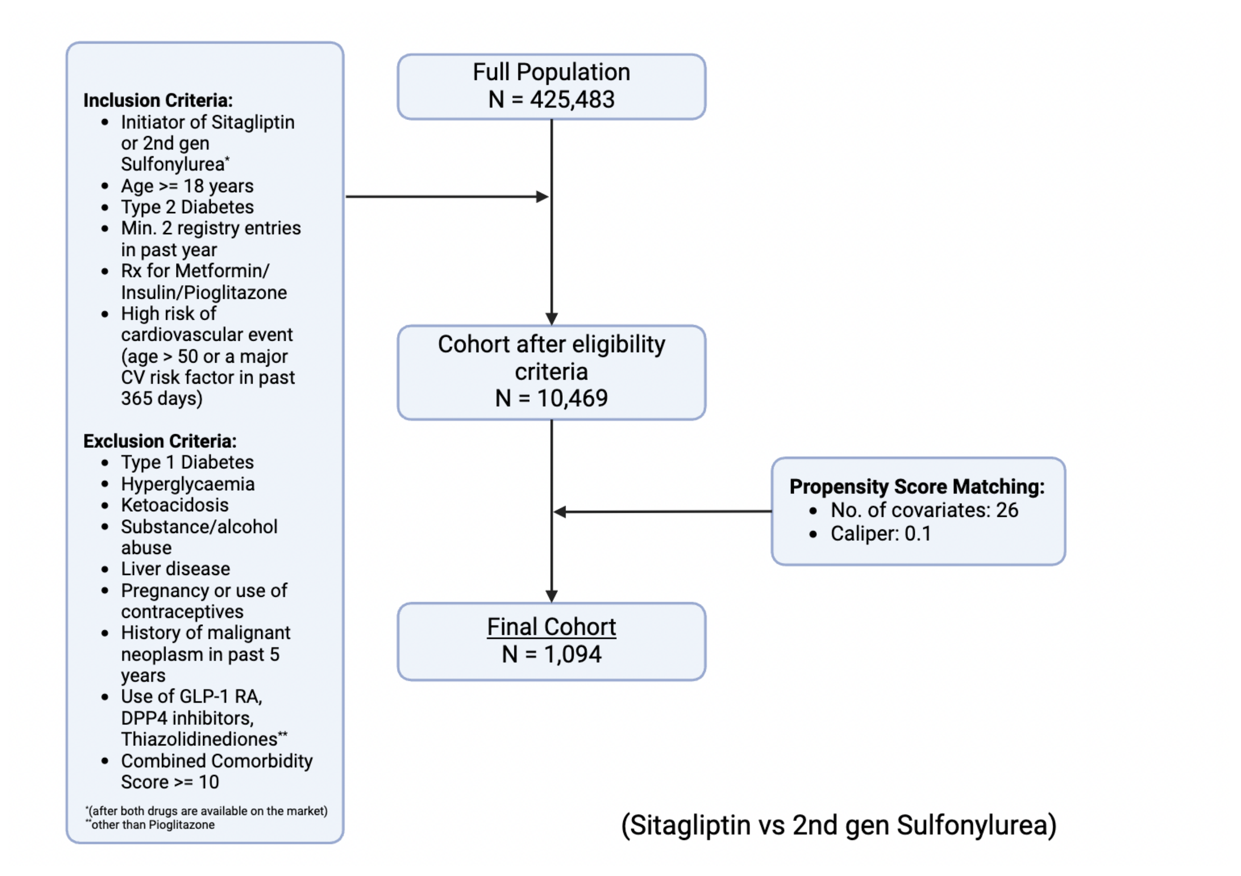


**Supplementary Figure 9. Cohort composition flowchart for Tecos trial emulation**
This flowchart illustrates the sample size changes along the incorporation of the eligibility (inclusion/exclusion) criteria and propensity score matching during the trial emulation.
A flowchart of the cohort formation can be found in **Supplementary Table 5**.
Details on the covariates used for the propensity score matching can be found in **Supplementary Table 9**.

### Supplementary Figure 10 – Cohort composition flowchart for Aristotle trial emulation

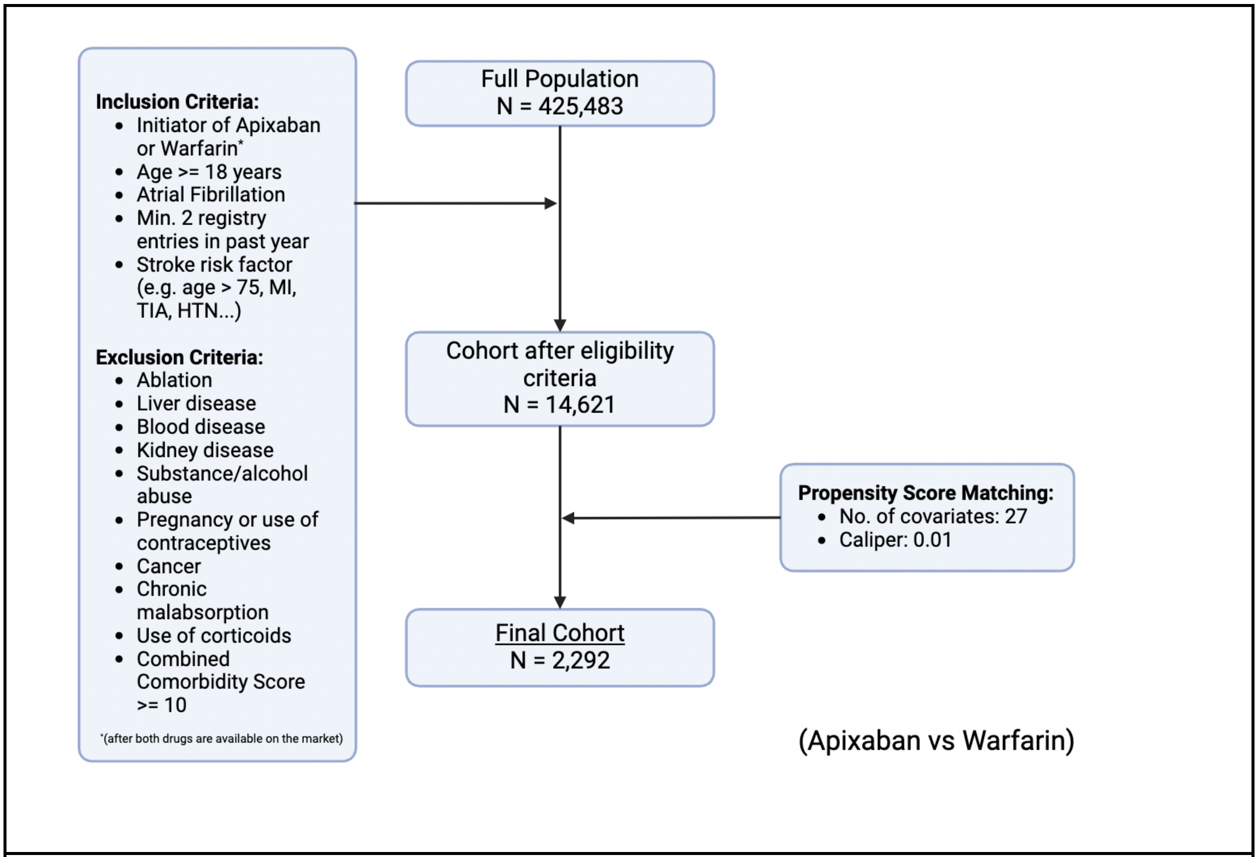


**Supplementary Figure 10. Cohort composition flowchart for Aristotle trial emulation**
This flowchart illustrates the sample size changes along the incorporation of the eligibility (inclusion/exclusion) criteria and propensity score matching during the trial emulation.
A flowchart of the cohort formation can be found in **Supplementary Table 6**.
Details on the covariates used for the propensity score matching can be found in **Supplementary Table 10**.

### Supplementary Figure 11 – Cohort composition flowchart for Rocket trial emulation

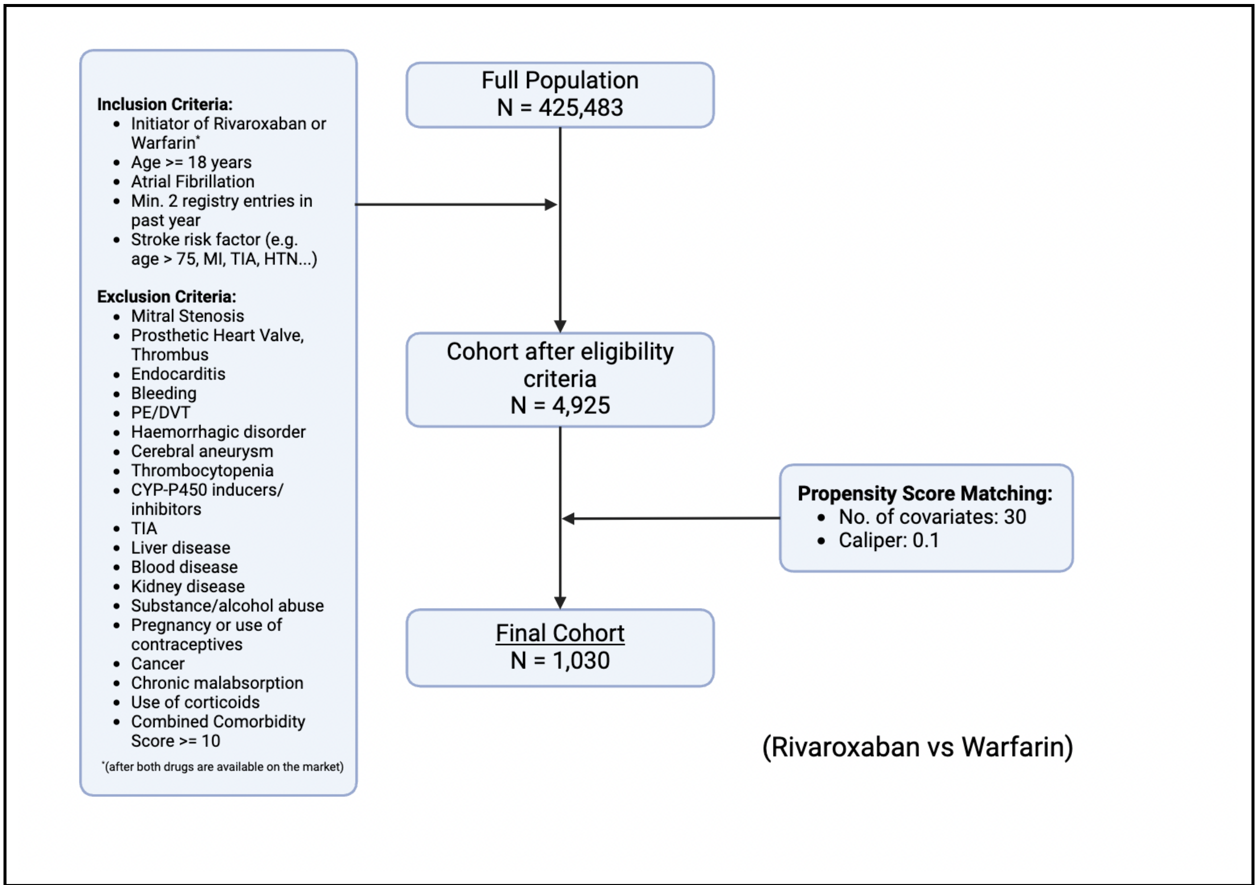


**Supplementary Figure 11. Cohort composition flowchart for Rocket trial emulation**
This flowchart illustrates the sample size changes along the incorporation of the eligibility (inclusion/exclusion) criteria and propensity score matching during the trial emulation.
A flowchart of the cohort formation can be found in **Supplementary Table 7**.
Details on the covariates used for the propensity score matching can be found in **Supplementary Table 11**.
